# Hierarchical representation learning of preeclampsia interactome connecting endometrial maturation, placentation, chorioamnionitis, and HELLP syndrome

**DOI:** 10.1101/2025.04.07.25325355

**Authors:** Herdiantri Sufriyana, Yu-Wei Wu, Emily Chia-Yu Su

## Abstract

**Background:** Existing proposed pathogenesis for preeclampsia (PE) was only applied for early-onset PE (EOPE). Our previous work identified the transcriptome to decipher EOPE and late-onset PE (LOPE), but the pathogenesis models were not validated. We developed and validated the pathogenesis models by hierarchical representation learning of interactomes connecting endometrial maturation, placentation, chorioamnionitis, and hemolysis, elevated liver enzyme, and low platelet (HELLP) syndrome.

**Methods:** We utilized 19 gene sets from our previous work to infer interactomes to develop (*n*=177) and validate (*n*=352) explainable artificial intelligence models for each PE subtype using deep-insight visible neural network and gradient-weighted class activation mapping.

**Results:** The hierarchically learned representations identified novel genes for LOPE, similar to endometrial maturation (*MRPL34, DYNLL1*), chorioamnionitis (*ANKRD13A*, *SLA*), and HELLP syndrome (*FAM43A*). We also identified novel genes for EOPE, similar to endometrial maturation (*SNAP23*, *PPL*, *LRRC32*), placentation (*GPT2*, *UBE2H*, *NIPAL3*, *NIN*, *KIAA0232*, *MT1F*, *DKK3*, *SLC24A3*), and HELLP syndrome (*SWAP70*, *GREM2*, *GPR146*, *PIP5K1B*, *EZR*). Nonetheless, a gene for each subtype was frequently studied, i.e., IGF1 (chorioamnionitis) and PAPPA2 (placentation), including LOPE and EOPE samples.

**Conclusions:** Our pathogenesis models connected both endometrial maturation and HELLP syndrome with LOPE and EOPE. However, they were differently connected with chorioamnionitis and placentation.

## Introduction

The two-stage dysfunction theory of preeclampsia (PE) has gained wide acceptance in explaining the pathogenesis of PE [1]. It belongs to two groups of pregnancy conditions that share similar pathophysiological derangements. The first group is pregnancy-induced hypertension (PIH) encompasses pre-existing chronic hypertension, gestational hypertension, and superimposed PE [2]. Meanwhile, the second group is placental dysfunction-related diseases (PDDs) or great obstetrical syndromes, which cover conditions PE with fetal growth restriction (FGR), isolated FGR, preterm labor with or without premature rupture of membranes, placental abruption, and second-trimester abortion [3, 4]. Moreover, PE may manifest towards the end of pregnancy before and after 34 weeks’ gestation respectively as early-onset PE (EOPE) and late-onset PE (LOPE) without or with clinical manifestation [5]. EOPE and LOPE are increasingly recognized for their distinct pathophysiological signatures [6–9]. Additionally, PE may result in hemolysis, elevated liver enzymes, and low platelet (HELLP) syndrome, although HELLP syndrome can occur independently of or precede PE [10]. However, the two-stage theory is specifically typical for and shared between EOPE and FGR.

The prevalence of PE ranges from 2.7 to 8.2% worldwide [11, 12]. LOPE accounts for 87.74% of PE cases among all singleton deliveries in Washington State, United States, from 2003 to 2008, with a prevalence of 2.72% for LOPE compared to 0.38% for EOPE [13]. Although EOPE is more severe than LOPE, it results in substantially poorer maternal and neonatal outcomes and leads to significant impact due to greater number of LOPE cases. LOPE resulted in a maternal mortality rate 2.67 times higher than that among women without PE (11.2 vs. 4.2 per 100,000 deliveries) and a 1.7 times higher rate of maternal morbidity, excluding obstetric trauma (95% confidence interval [CI] 1.6, 1.9) [14]. Meanwhile, perinatal mortality and morbidity in LOPE were twice as high as those from normotensive pregnant women (95% CI 1.8, 2.3) [13, 15]. Therefore, exploring the pathogenesis beyond the two-stage theory is expected to deepen our understanding of PE.

This PE pathogenesis theory proposes placental dysfunction in the first trimester, occurring before endothelial dysfunction that leads to clinical manifestations [1]. The antecedents of placental dysfunction remain largely unknown. Several possibilities have been revealed through associations between PE and endometrial maturation or decidualization [16–20], metagenomics profiling of the placenta [21–24], and maternal susceptibility to vascular dysfunction [25, 26]. Differentially expressed genes (DEGs) significantly overlapped between those from preeclamptic chorionic villi sampling (CVS) and those from pathological endometrium [16]; however, the overlapped DEGs were co-expressed rather than counter-expressed, indicating co-occurrence rather than a potential causal-effect relationship. Regarding the placental microbiome and PE, although 57 studies identified a low biomass microbiota in placenta [23], previous publications on the role of placental microbiota in PE pathogenesis mostly consist of proof-of-concept reviews [21, 22]. Oppositely, another study suggested that bacterial and viral abundance in the placenta might play a limited role in the pathogenesis of PE [24]. However, their method only encompassed 675 bacterial species and 20 viral species, utilizing a technique based on sequencing small non-coding RNA, which increases the microorganism-to-host ratio. Related to maternal vascular susceptibility [26], a genome-wide association study identified genes associated with the blood pressure trait in PE, exhibiting pleiotropic effects on cardiometabolic, endothelial, and placental functions. However, none of these studies have specifically identified human biological targets linked to the antecedents of placental dysfunction that distinguishes between EOPE and LOPE.

Previously, we identified transcriptomes that may explain a universal PE pathogenesis [27]. We proposed that deranged endometrialization, but not decidualization, may lead to placental dysfunction resulting in EOPE; if placental dysfunction does not occur, the result is LOPE instead. Gene regulation was also proposed to pivot the microbial composition in the endometrium, switching between PE and chorioamnionitis in opposite direction. Nevertheless, transcriptome data alone cannot elucidate the pathogenesis of PE unless we develop pathogenesis models using the identified transcriptome. The modeling of pathogenesis must facilitate validation with an independent dataset and incorporate inter-gene relationships in addition to their correlations with a specific PE subtype. These objectives could be achieved through predictive modeling via hierarchical representation learning, i.e., a deep insight visible neural network (DI-VNN) [28]. We aimed to develop and validate pathogenesis models of PE subtypes by hierarchical representation learning of their interactomes connecting endometrial maturation, decidualization, placentation, chorioamnionitis, and HELLP syndrome.

## Methods

### Study design

We utilized gene sets which were discovered in our previous work [27] to infer interactomes for each PE subtype. An interactome was subsequently used to construct an XAI model for each of pathogenesis models of a PE subtype. We developed and externally validated these models using different microarray datasets of mRNA expression. They were retrieved from two publicly-accessed databases, i.e., Gene Expression Omnibus (GEO) and ArrayExpress. Predictive modeling was applied as an approach to justify a pathogenesis model in this study. A sample was predicted to be negative or positive of a PE subtype (i.e., an outcome) by a gene set (i.e., predictors) in an XAI model. Ethical review was exempted by the Taipei Medical University Joint Institutional Review Board (TMU-JIRB number: N202106025).

The datasets consisted of mRNA expression data of total RNA extracts. Three datasets with the same microarray platform were utilized for the model development set (*n*=177), while five datasets with diverse microarray platform were utilized for the model test set (*n*=352). No samples in the test set were the same with those in the development set. Neither was the same with those in our previous work, which was used to discover the gene sets for the pathogenesis models. The test set was also normalized using the quantile distribution of controls in the development set. Details on data preprocessing and gene set discovery were described in our previous work [27]. Therefore, the test set is independent and challenging. The latter was contributed by its diverse biological and technical characteristics which facilitated a robust test for our pathogenesis models.

For outcome definition, samples were positives if they were: (1) late-onset, severe PE; (2) early-onset, severe PE; (3) late-onset, mild-to-moderate PE with FGR; and (4) late-onset, severe PE with FGR. Meanwhile, the samples were negatives if they were normotension without FGR. These outcome definitions were initially determined by combining four axes related to PE: (1) PIH; (2) PDDs; (3) onset; and (4) severity. However, considering sample size estimation and gene set discovery, both of the development and test sets were only sufficient for 4 of 28 potential combinations. Details on outcome definition and sample size estimation were described in our previous work [27].

### Hierarchical representation learning

For predictors in the pathogenesis models (Table 1, Table S1), we used gene sets which were discovered in our previous work [27]. A gene set was selected if it was a significant overlap between a pair of DEGs of a PE subtype and each of the associated factors. For the gene sets in this study, the associated factors were: (1) endometrial maturation; (2) placentation; (3) chorioamnionitis; and (4) HELLP syndrome. The DEGs of a PE subtype were identified by comparing normotension without FGR with each subtype. For endometrial maturation, the DEGs were identified by comparing proliferative-with early-, mid-, or late-secretory endometrium. Meanwhile, the DEGs of placentation were identified by comparing term with either first- or second-trimester placenta. We also identified the placental DEGs by comparing samples without and with either chorioamnionitis or HELLP syndrome at second and third trimesters. However, the gene set of HELLP syndrome at second trimester did not pass the selection criterion. Eventually, none of samples from the test set was used to determine these gene sets. Hereafter, we shall not mention the control groups of the gene sets and the term “overlapped DEGs” for brevity.

**Table 1.** Predictors in the pathogenesis models.

| Representation of |  | Number of genes |  |  |  |
| --- | --- | --- | --- | --- | --- |
|  |  | PE |  | PE with FGR |  |
|  |  | Late-onset,<br>severe | Early-onset,<br>severe | Late-onset,<br>mild-to-moderate | Late-onset,<br>severe |
| Endometrial maturation | Early-secretory phase | 0 | 468 | 37 | 360 |
|  | Mid-secretory phase | 72 | 0 | 0 | 0 |
|  | Late-secretory phase | 58 | 405 | 0 | 304 |
| Placentation | First trimester | 0 | 353 | 0 | 280 |
|  | Second trimester | 0 | 44 | 0 | 41 |
| Chorioamnionitis | Second trimester | 0 | 24 | 0 | 20 |
|  | Third trimester | 58 | 426 | 0 | 375 |
| HELLP syndrome | Third trimester | 52 | 283 | 0 | 189 |

FGR, fetal growth restriction; HELLP, hemolysis, elevated liver enzyme, and low platelet; PE, preeclampsia.

Interactomes were identified from for each gene set by knowledge- and data-driven ontology inference methods. The former applied pathway enrichment analysis (PEA) utilizing gene ontology (GO) database, while the latter applied clique-extracted ontology (CliXO) algorithm. Both resulted in directed acyclic graphs (DAGs) which would be used for constructing neural network architectures of the XAI models. Therefore, there were potentially 38 pathogenesis models from 19 gene sets and 2 ontology inference methods. However, no ontology might be inferred by an ontology inference method for a gene set.

For PEA utilizing GO database, we identified GO terms in two phases. First, we selected GO terms of which genes were enriched by DEGs of each of the associated factors. Second, among the selected GO terms for each of the associated factors, we identified those of which genes were enriched by each gene set in the pathogenesis models. Before any phases, GO terms were excluded if the evidence codes were IEA and IGI, respectively for limiting only human-curated terms [29] and acyclic graphs [30] for the neural network architectures. We conducted each enrichment analysis separately among GO terms with 15–200, 201–500, 501–2000 and >2000 genes, to avoid irrelevant stringency in multiple testing correction due to gene abundance [29]. All the significantly enriched child terms were connected to the next ancestors up to the root if the parent term was not significantly enriched. We constructed a DAG using GO with only significantly enriched terms for each pathogenesis model.

For CliXO algorithm, we inferred ontologies using each gene set in the pathogenesis models. This algorithm finds maximal cliques in a gene network by progressive thresholding of the similarity scores (i.e., Pearson correlation coefficients). We added a root ontology to be the parent of the inferred ontologies at the most ancestor positions. In addition, we aligned the inferred ontologies with GO using network-extracted (NeXO) algorithm for assisting the ontology interpretation. We considered an ontology were novel if it was not significantly aligned to any GO terms.

We developed and externally validated the XAI models applying a DI-VNN [28]. The input representation of DI-VNN was a three-dimensional feature map onto which we mapped gene sets. The coordinates were determined by *t*-moderated stochastic neighbor embedding (*t*-SNE) algorithm with Barnes-Hut approximation. This algorithm mapped genes according to the similarity scores which were also used for CliXO algorithm. For example, if we have 468 genes, then there are 468 pixels on the feature map. Different gene sets had different feature maps. However, all samples used the same feature map for each gene set, while the gene expression values were different among samples. We transformed the expression values of each gene by 1-bit stochastic gradient descent into 1 and -1, if they were respectively ≥ and < the average, resembling up- and down-regulated gene expression at individual level. Zero values were assigned to any coordinates in a feature map, that were unoccupied by any genes.

Using each interactome, we subset a feature map hierarchically by the inferred ontologies. For example, if an ontology included 2 of 468 genes, then there are only 2 pixels on the 468-gene, feature map that was subset by that ontology. Specifically, we assigned zero values for the remaining 466 genes for any samples. Each subset predicted an outcome; thus, we had multiple versions of an outcome prediction as many as the number of the inferred ontologies for each pathogenesis model. We also obtained a learned representation for each ontology. Both prediction and learned representation of a parent ontology also included the inputs and the learned representations of its children ontologies. Eventually, to train or fit the DI-VNNs, we followed the same hyperparameters as the previous study [28]. Briefly, a pathogenesis model was trained as a single network consisting multiple gene subsets based on the inferred ontologies. The predicted probabilities of the root and non-root ontologies contributed the losses with weights of 1 and 0.3.

### Statistical analysis

To evaluate the pathogenesis models, we used the test sets to compute the average values and 95% confidence intervals (CIs) of positive predictive value (PPV), true positive rate (TPR), negative predictive value (NPV), and true negative rate (TPR). The interval estimates were obtained by 30-time bootstrapping of each test set. A threshold of the predicted probabilities was chosen based on the development set if the threshold resulted in maximum difference of net benefits with those of the references. They were treat-all and treat-none scenarios in which all samples were respectively predicted as positive and negative. A pathogenesis model was valid at ontology level if there was at least 1 ontology of which the lower bounds were finite and >0 for a pair of evaluation metrics, i.e.: (1) PPV-TPR; or (2) NPV-TNR. These criteria should be fulfilled using both development and test sets. Respectively, the pathogenesis models were valid for representing positives or negatives of an outcome. Furthermore, a pathogenesis model was valid at ontology level if at least 1 ontology had an absolute difference of ≤25% between the averages using the development and test sets for the pair of evaluation metrics. We also evaluate explainability of the valid pathogenesis models using gradient-weighted class activation mapping (grad-CAM) method. A pathogenesis model was valid at gene level if at least 1 gene has a grad-CAM value >97.5^th^ percentile of those in the same ontology. Eventually, we used gene set artificial intelligence (GSAI), which was powered by a large language model (LLM) [31], to annotate the function of the genes for each ontology of a pathogenesis model. We selected GPT-4 as an LLM which was used for GSAI. The up- and down-regulation status of the genes from our previous study [27] were also incorporated in the GSAI response summary.

## Results

### Pathogenesis models

Of 19 gene sets, we identified 9 interactomes (47%, *n*=9/19) by PEA utilizing GO database and 19 interactomes (100%, *n*=19/19) by CliXO algorithm (Figure 1). All the 28 interactomes resulted in 28 pathogenesis models. Only 10 (36%, *n*=10/28) and 7 (70%, *n*=7/10) of them fulfilled the criteria respectively at ontology and gene levels. Eventually, seven pathogenesis models were validated for 2 of 4 PE subtypes, i.e., late- and early-onset, severe PE without FGR. Hereafter, we only mention LOPE and EOPE for brevity. Both are severe and isolated (i.e., without FGR) in this study. The sample characteristics were already described in our previous study [27].

**Figure 1.**
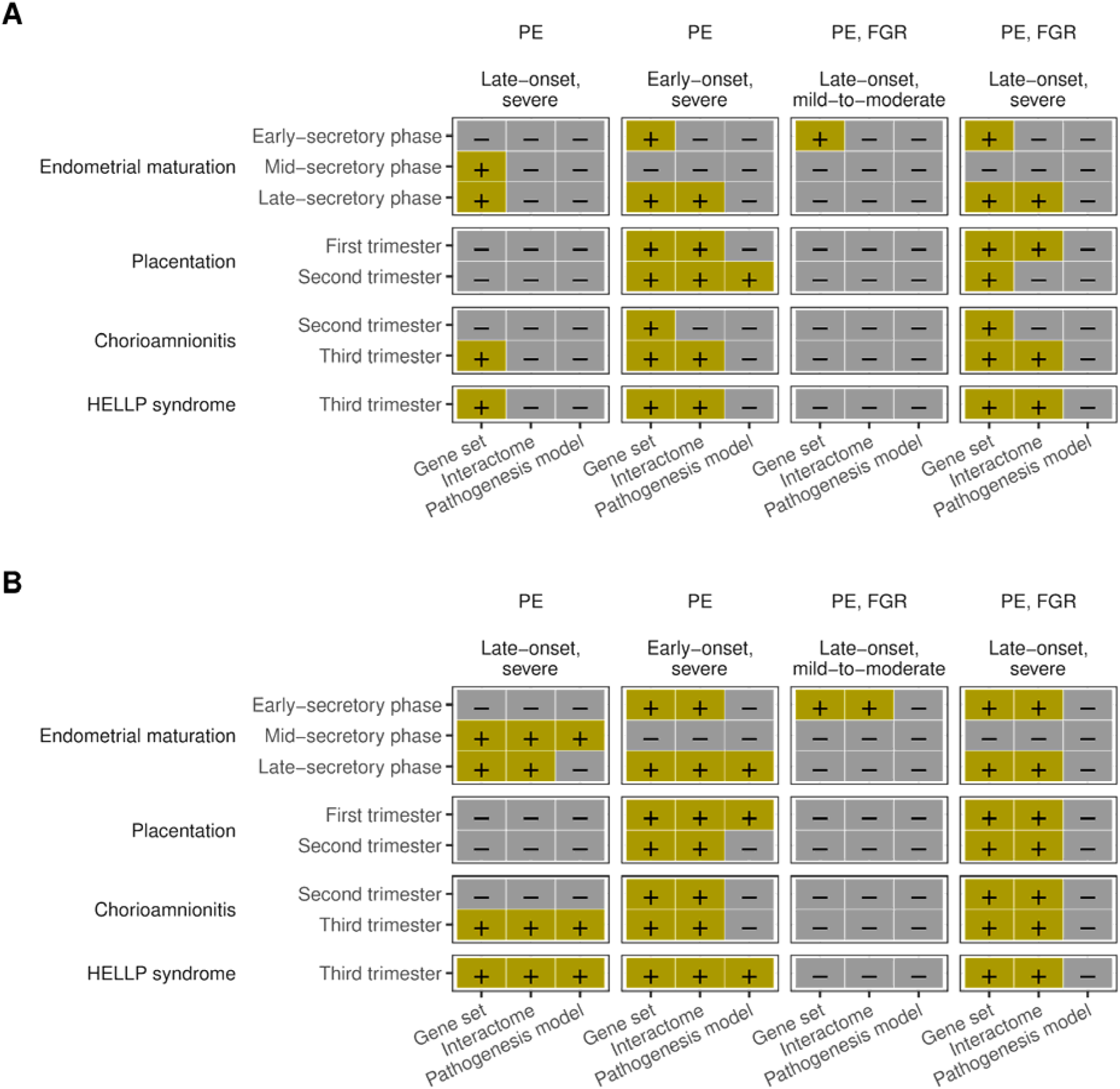
Availability of gene sets, interactomes, and pathogenesis models of a PE subtype and each of the associated factors. Positive (+) and negative (-) symbols respectively mean “available” and “not available”. Colors are assigned to improve visualization by augmenting the symbols. FGR, fetal growth restriction; HELLP, hemolysis, elevated liver enzyme, and low platelet; PE, preeclampsia.

All the pathogenesis models for LOPE fulfilled the criteria at ontology level for PPV and TPR; thus, they directly represented a positive outcome. For EOPE, the pathogenesis models fulfilled the criteria at ontology level for NPV and TNR; thus, they represented a negative outcome. If the prediction was negative, the outcome was truly normotension without FGR. However, the outcome might or might not be EOPE, if the prediction was positive. Other factors were also needed to sufficiently result in EOPE.

### Hierarchical learning representation of late-onset, severe preeclampsia

Three pathogenesis models were valid for LOPE. The gene sets were overlaps between DEGs of LOPE and these associated factors: (1) mid-secretory endometrium (Figure 2A); (2) third-trimester placenta under chorioamnionitis (Figure 2B); and (3) third-trimester placenta under HELLP syndrome (Figure 2C). Respectively, the functions of the eligible gene sets were annotated by GSAI as (Table S2): (1) “Mitochondrial Ribosome Biogenesis and Cellular Respiration” with confidence score of 0.85; (2) “Regulation of Protein Ubiquitination and Circadian Rhythm”, “Protein Ubiquitination and Immune Response Regulation”, and “Insulin-like Growth Factor Signaling” respectively with confidence scores of 0.85, 0.85, and 1.00; and (3) “Cyclic Guanosine Monophosphate (cGMP) Degradation” with confidence score of 0.95.

**Figure 2.**
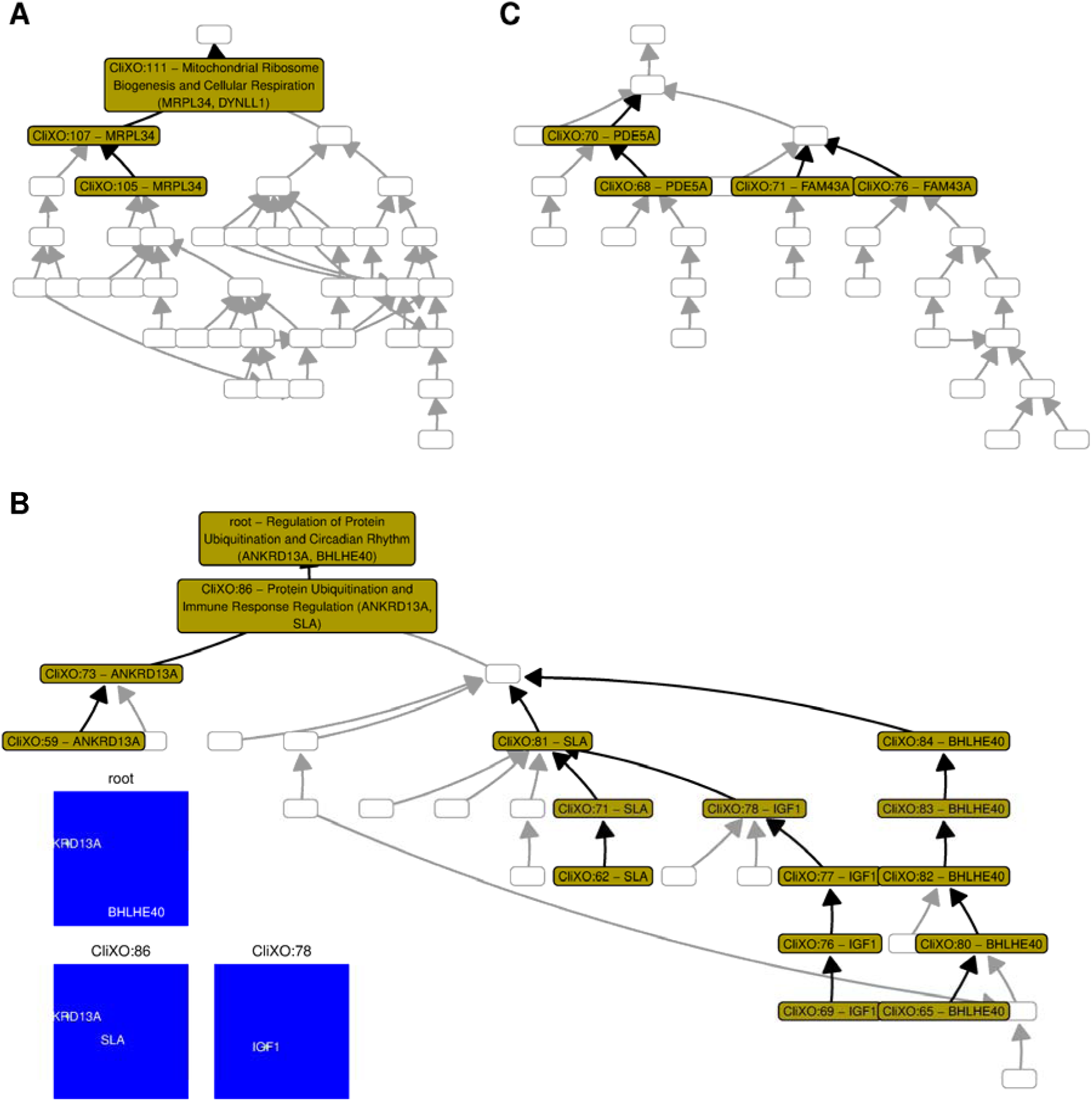
Hierarchical learning representation of late-onset, severe PE interactome connecting: (A) mid-secretory endometrium; (B) third-trimester placenta under chorioamnionitis; and (C) third-trimester placenta under HELLP syndrome. Learning representations are shown for three ontologies in the pathogenesis model B to show how gradients (i.e., non-blue colors) shift on different genes across these ontologies. A node for an ontology is colored and labeled if the ontology was eligible and has at least 1 gene that was eligible. A label consists of ontology code and name with gene names (i.e., HGNC symbols). If there is only 1 gene, no ontology name is shown. To improve path visualization, an edge from a colored node is made bolder than that from uncolored node. CliXO, clique-extracted ontology; HELLP, hemolysis, elevated liver enzyme, and low platelet; HGNC, the human genome organization gene nomenclature; PE, preeclampsia.

In mid-secretory endometrium, two genes were downregulated compared to proliferative endometrium, i.e., *MRPL34* and *DYNLL1* (Table 2). Based on GSAI annotation, the former protein is a part of mitochondrial ribosome which synthesizes proteins primarily involved in cellular ATP production, while the latter protein interacts with the outer membrane of mitochondria in regulating its morphology and function (Table S2). However, they were upregulated in third-trimester placenta under LOPE. In the pathogenesis model (Figure 2A), *MRPL34* expression alone was predictive for LOPE. Including *DYNLL1* in the same ontology (i.e., CliXO:111) with *MRPL34* did not change its predictive role.

**Table 2.** Discovery of gene set function by GSAI for late-onset, severe PE.

| Representation of |  | Ontology | GSAI<br>Query | Response summary* |
| --- | --- | --- | --- | --- |
| Endometrial maturation | Mid-secretory phase | CliXO:111 | MRPL34, DYNLL1 | The proteins (i.e., MRPL34 [D/U] and DYNLL1 [D/U] †) in this system are involved in the biogenesis of the mitochondrial ribosome and the regulation of mitochondrial function, both of which are essential for cellular respiration. The interplay between these proteins likely ensures the efficient production of ATP, the cell's main energy currency. The synergistic effect of these proteins contributes to the maintenance of cellular energy homeostasis. |
| Chorio-amnionitis | Third trimester | root | ANKRD13A, BHLHE40 | The proteins in this system are involved in the regulation of protein ubiquitination and circadian rhythm. ANKRD13A [D/D] contributes to the process of protein ubiquitination, which is crucial for protein degradation and cellular localization. BHLHE40 [D/U], on the other hand, is involved in the regulation of circadian rhythm, contributing to the maintenance of the body's internal clock. The interplay between these proteins could potentially influence the balance between protein degradation and synthesis, as well as the synchronization of cellular processes with the circadian rhythm. |
|  |  | CliXO:86 | ANKRD13A, SLA | The proteins in this system are primarily involved in the regulation of protein ubiquitination and immune response. ANKRD13A [D/D] contributes to the maintenance of protein homeostasis through its role in protein ubiquitination and degradation, while SLA [U/U] plays a crucial role in immune response regulation through its inhibitory effect on TCR signaling. These processes are essential for maintaining cellular function and immune system balance. |
|  |  | CliXO:78 | IGF1 | The IGF1 [U/D] protein is a key player in the IGF signaling system, which is a complex network of interactions that regulate cell growth, differentiation, and survival. The system exhibits both synergistic and antagonistic effects, depending on the context and the specific interactions involved. |
| HELLP syndrome | Third trimester | CliXO:70 | PDE5A | The primary biological process performed by PDE5A [D/D] is the degradation of cGMP, a process that has significant implications for vascular and neuronal function. The protein's role in this process is well-established, hence the high confidence score. |
|  |  | CliXO:71 | FAM43A | Without additional proteins for analysis, the system can only be identified as a "System of unrelated proteins" with a confidence score of 0.00. Further research and data are needed to elucidate the specific biological processes and interactions involving FAM43A [U/U]. |

\*, see Table S2 for response details and each mention of gene name is modified by including up (U)- or down (D)-regulation status of respectively the associated factor and third-trimester placenta under late-onset, severe PE; †, we redacted a part in parentheses to mention gene names, that were related to that part based on Table S2, if they were not mentioned in the response summary; CliXO, clique-extracted ontology; GSAI, gene set artificial intelligence; HELLP, hemolysis, elevated liver enzyme, and low platelet; PE, preeclampsia.

In third-trimester placenta under chorioamnionitis, *IGF1* was upregulated (Table 2). IGF1 was annotated by GSAI as a protein that activates IGF1R in regulation of cell growth, differentiation, and survival (Table S2). However, *IGF1* were downregulated in third-trimester placenta under LOPE. In the pathogenesis model (Figure 2B), *IGF1* expression alone was predictive for LOPE, but it was less predictive in CliXO:81 which also included *SLA*. It was upregulated in third-trimester placenta under either chorioamnionitis or LOPE (Table 2). SLA was annotated by GSAI as a negative regulator of T-cell receptor signaling, which in turns, prevent overactive immune responses (Table S2).

Furthermore, our pathogenesis model put larger weights on *ANKRD13A* expression than those on *SLA* and *BHLHE40* if they were included in the same ontologies, i.e., CliXO:86 and root, respectively (Figure 2B). *ANKRD13A* expression alone was predictive for LOPE. ANKRD13A was annotated by GSAI to be involved in ubiquitination which regulates protein degradation (Table S2). However, *ANKRD13A* was downregulated in third-trimester placenta under either chorioamnionitis or LOPE (Table 2). The difference was that *BHLHE40* was downregulated in third-trimester placenta under chorioamnionitis but upregulated in that under LOPE. GSAI annotated BHLHE40 as playing a role in (Table S2): (1) regulation of circadian rhythm and (2) regulation of gene expression in response to hypoxia. We considered the latter was more relevant in this study based on the tissue context, i.e., placenta. In the pathogenesis model (Figure 2B), *BHLHE40* expression alone was also predictive for LOPE.

In third-trimester placenta under HELLP syndrome, *PDE5A* and *FAM43A* were respectively downregulated and upregulated (Table 2). The gene expressions were regulated the same way for LOPE. Either *PDE5A* or *FAM43A* alone was also predictive for LOPE but neither if both genes were included in the same ontology (Figure 2C). GSAI annotated PDE5A as playing a role in (Table S2): (1) regulation of vascular tone and (2) regulation of neuronal signaling. We considered the former was more relevant in this study based on the tissue context, i.e., placenta. Downregulation of *PDE5A* may resemble its inhibition by sildenafil, as annotated by GSAI, i.e., enhancing nitric oxide on its vasodilation effect, hence, decreasing blood pressure and improving blood flow. Unfortunately, since a role of FAM43A in cellular processes depends on other proteins it interacts with, it could not be annotated by GSAI without information of the other proteins (Table 2).

### Hierarchical learning representation of early-onset, severe preeclampsia

Four pathogenesis models were valid for EOPE. The gene sets were the overlaps between DEGs of EOPE and these associated factors: (1) late-secretory endometrium (Figure 3A); (2) first-trimester placenta in which the interactome was inferred by CliXO algorithm (Figure 3B); (3) second-trimester placenta in which the interactome was inferred by PEA using GO database (Figure 3C); and (4) third-trimester placenta under HELLP syndrome (Figure 3D). Respectively, the functions of the eligible gene sets were annotated by GSAI as (Table S3): (1) “Lipid Metabolism and Cellular Adhesion” with confidence score of 0.85; (2) “Glutamate Metabolism and Protein Ubiquitination”, “Mitochondrial Metabolism and Neurodevelopment”, and “Regulation of Immune Response and Neuronal Survival” respectively with confidence scores of 0.85, 0.85, and 0.85; (3) “Regulation of Extracellular Matrix Remodeling and Ion Transport” with confidence score of 0.85; and (4) “Regulation of Cellular Differentiation and Signal Transduction” with confidence score of 0.85.

**Figure 3.**
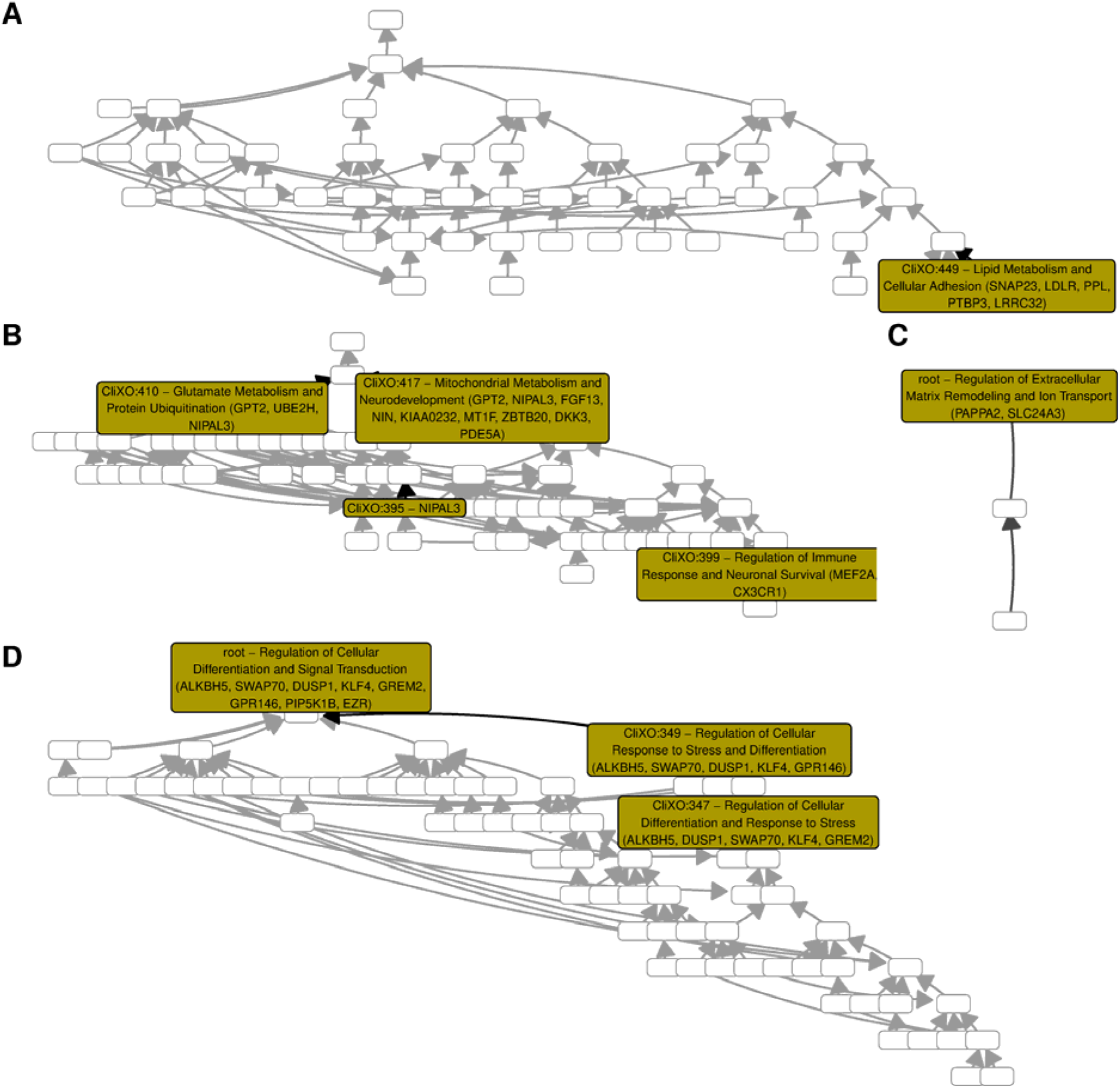
Hierarchical learning representation of early-onset, severe PE interactome connecting: (A) late-secretory endometrium; (B) first-trimester placenta in which interactome was inferred by CliXO algorithm; (C) second-trimester placenta in which interactomes were inferred by PEA using GO database; and (D) third-trimester placenta under HELLP syndrome. A node for an ontology is colored and labeled if the ontology was eligible and has at least 1 gene that was eligible. A label consists of ontology code and name with gene names (i.e., HGNC symbols). If there is only 1 gene, no ontology name is shown. To improve path visualization, an edge from a colored node is made bolder than that from uncolored node. CliXO, clique-extracted ontology; GO, gene ontology; HELLP, hemolysis, elevated liver enzyme, and low platelet; HGNC, the human genome organization gene nomenclature; PE, preeclampsia; PEA, pathway enrichment analysis.

In late-secretory endometrium, five genes were upregulated (Table 3). All the genes were also upregulated in third-trimester placenta under LOPE, except *SNAP23* which was downregulated. Based on GSAI annotation, this protein is involved in vesicle docking and fusion to target membranes for secreting lipoproteins or other molecules (Table S3). The pathogenesis model required all the five gene expressions to be predictive (Figure 3A). Downregulation of *SNAP23* and upregulation of the remaining genes might or might not result in EOPE, but the opposite regulation resulted in normotension without FGR. As implied by GSAI annotation (Table S3), the opposite regulation of *SNAP23*, *LDLR*, and *PTBP3* may respectively lead to: (1) more secretion of lipoproteins and other molecules; (2) less endocytosis of low-density lipoprotein into cells; and (3) less alternative splicing related to lipid metabolism. Meanwhile, the opposite regulation of *PPL* and *LRRC32* may lead to less cellular resistance to mechanical stress via cell adhesion. The latter may also lead to less suppression of the immune response via regulatory T cells.

**Table 3.** Discovery of gene set function by GSAI for early-onset, severe PE.

| Representation of |  | Ontology | GSAI<br>Query | Response summary* |
| --- | --- | --- | --- | --- |
| Endometrial maturation | Late-secretory phase | CliXO:449 | SNAP23, LDLR, PPL, PTBP3, LRRC32 | The proteins in this system are involved in lipid metabolism (i.e., SNAP23 [U/D], LDLR [U/U], and PTBP3 [U/U] †), cellular adhesion (i.e., PPL [U/U] and LRRC32 [U/U] †), and the regulation of the immune response (i.e., LRRC32 [U/U] †). The interplay between these proteins allows for the regulation of cholesterol levels, the maintenance of cellular structure and integrity, and the modulation of the immune response. The proteins in this system work together to ensure the proper functioning of these processes, with potential synergistic effects in lipid metabolism and cellular adhesion. |
| Placentation | First trimester | CliXO:410 | GPT2, UBE2H, NIPAL3 | The proteins in this system are primarily involved in the metabolism of amino acids, protein ubiquitination, and potentially lipid metabolism. These processes are interconnected, as the degradation of proteins (mediated by UBE2H [U/D]) can provide amino acids for further metabolism (mediated by GPT2 [U/U]), and both processes could potentially interact with lipid metabolism (mediated by NIPAL3 [U/D]). The interplay between these proteins and their associated processes contributes to the maintenance of cellular homeostasis. |
|  |  | CliXO:417 | GPT2, NIPAL3, FGF13, NIN, KIAA0232, MT1F, ZBTB20, DKK3, PDE5A | The proteins in this system are primarily involved in mitochondrial metabolism (i.e., GPT2 [U/U] and NIPAL3 [U/D] †) and neurodevelopment (i.e., FGF13 [D/D], NIN [D/D], and ZBTB20 [D/D] †), with some proteins also contributing to cellular detoxification (i.e., MT1F [U/U] †), embryonic development (i.e., DKK3 [D/D] †), and vascular regulation (i.e., PDE5A [D/D] †). The interplay between these proteins likely contributes to the overall function of the system, with potential synergistic effects in energy metabolism and neurodevelopment, and antagonistic effects in cellular detoxification and development. (KIAA0232 [D/D] is predicted to be a membrane protein involved in cellular transport processes. †) |
|  |  | CliXO:399 | MEF2A, CX3CR1 | The interaction between MEF2A [D/U] and CX3CR1 [D/D] suggests a complex interplay between the immune system and the nervous system. MEF2A [D/U], through its role in gene regulation, could potentially influence the expression of CX3CR1 [D/D] and thus modulate immune responses. Conversely, signals received through CX3CR1 [D/D] could impact the activity of MEF2A [D/U], influencing neuronal survival and function. This system of interacting proteins, therefore, appears to be involved in the regulation of immune responses and neuronal survival, with potential implications for diseases involving neuroinflammation and neurodegeneration. |
|  | Second trimester | root | PAPPA2, SLC24A3 | The proteins in this system are involved in the regulation of extracellular matrix remodeling and ion transport. PAPPA2 [D/U]'s role in ECM remodeling can influence cell behavior and tissue structure, while SLC24A3 [U/U]'s role in ion transport can affect various cellular processes. These proteins may interact in a way that their functions complement each other, contributing to the overall homeostasis of the cell. |
| HELLP syndrome | Third trimester | root | ALKBH5, SWAP70, DUSP1, KLF4, GREM2, GPR146, PIP5K1B, EZR | The proteins in this system are involved in various aspects of cellular differentiation and signal transduction. They contribute to the regulation of gene expression (i.e., ALKBH5 [U/U] †), actin cytoskeleton organization (i.e., SWAP70 [U/U] and PIP5K1B [D/D] †), MAP kinase signaling (i.e., DUSP1 [U/U] †), stem cell pluripotency (i.e., KLF4 [U/U] †), organogenesis (i.e., GREM2 [U/U] †), and potentially other signaling pathways (i.e., GPR146 [U/U] and EZR [U/U] †). Their interplay likely results in a complex network of regulatory mechanisms that control cellular differentiation and response to external signals. |

\*, see Table S3 for response details and each mention of gene name is modified by including up (U)- or down (D)-regulation status of respectively the associated factor and third-trimester placenta under early-onset, severe PE; †, we redacted a part in parentheses to mention gene names, that were related to that part based on Table S3, if they were not mentioned in the response summary by GSAI; CliXO, clique-extracted ontology; GSAI, gene set artificial intelligence; HELLP, hemolysis, elevated liver enzyme, and low platelet; PE, preeclampsia.

In first-trimester placenta, *NIPAL3* and *UBE2H* was upregulated (Table 3). However, both were downregulated in third-trimester placenta under EOPE. In the pathogenesis model (Figure 3B), *NIPAL3* expression alone was predictive. Downregulation of *NIPAL3* might or might not result in EOPE, but the upregulation resulted in normotension without FGR. As implied by GSAI annotation (Table S3), the upregulation of *NIPAL3* lead to the transport of magnesium across membranes, which is a critical cofactor for many enzymes.

Meanwhile, UBE2H was annotated by GSAI as one of ubiquitin-conjugating enzymes (Table S3). Its upregulation may lead to more removal of misfolded or damaged proteins. *UBE2H* expression were predictive if it was included in the same ontology with NIPAL3 and GPT2, i.e., CliXO:410 (Figure 3B). The latter was annotated by GSAI as an enzyme which converts alanine and alpha-ketoglutarate, provided by protein degradation via UBE2H, to pyruvate and glutamate.

However, CliXO:417 was predictive without *UBE2H* expression, in which *GPT2* and *NIPAL3* were predictive with other seven genes. These genes shared the same up- and down-regulation in: (1) first-trimester, or (2) third-trimester placenta under EOPE (Table 2). As implied by GSAI annotation (Table S3), the placental context of their regulations might or might not result in EOPE, but the opposite regulations resulted in normotension without FGR, leading to: (1) more cellular development (i.e., upregulation of *FGF13*, *NIN*, *ZBTB20*, and *DKK3*) and transport (i.e., upregulation of KIAA0232); (2) less cellular detoxification via metallothioneins which highly contains cysteine residues (i.e., downregulation of MT1F); and (3) less vasodilation effect of nitric oxide, hence, increasing blood pressure and reducing blood flow (i.e., upregulation of *PDE5A*). Nonetheless, including those genes in the same ontology (i.e., CliXO:417) with *NIPAL3* did not change its predictive role.

In the same pathogenesis model (Figure 3B), *MEF2A* and *CX3CR1* expressions were also predictive without the abovementioned genes. In first-trimester placenta, *MEF2A* and *CX3CR1* was downregulated (Table 3). However, *MEF2A* had the opposite regulation in third-trimester placenta under EOPE. Up- and down-regulations of respectively *MEF2A* and *CX3CR1* might or might not result in EOPE, but the opposite regulations resulted in normotension without FGR. As implied by GSAI annotation (Table S3), the downregulation of *MEF2A* lead to: (1) less transcription of genes involved in survival and differentiation of neurons and muscle cells, and synaptic plasticity, and (2) less transcription of genes involved in T-cell activation and differentiation. We considered the latter was more relevant in this study based on the tissue context, i.e., placenta. Meanwhile, the upregulation of *CX3CR1* lead to migration and adhesion of various immune cells, including monocytes, macrophages, T cells, and microglia.

In second-trimester placenta, *PAPPA2* and *SLC24A3* was respectively down- and upregulated (Table 3). However, *PAPPA2* had the opposite regulation in third-trimester placenta under EOPE compared to that under normotension without FGR. The pathogenesis model (Figure 3C) required both *PAPPA2* and *SLC24A3* to be predictive. Upregulations of *PAPPA2* and *SLC24A3* might or might not result in EOPE, but the opposite regulations resulted in normotension without FGR. As implied by GSAI annotation (Table S3), the downregulation of *PAPPA2* lead to: (1) less cleaving of IGFBP5, which in turn, reducing of released IGFs involved in cell growth, development, and metabolism; and (2) less remodeling of extracellular matrix. Meanwhile, the downregulation of *SLC24A3* lead to less exchange between sodium and calcium, that is potassium-dependent, to maintain their intracellular concentrations.

In third-trimester placenta under HELLP syndrome, at least four genes were upregulated (Table 3). These genes were: (1) *ALKBH5*; (2) *SWAP70*; (3) *DUSP1*; and (4) *KLF4*. They were also regulated the same way for EOPE. To be predictive in the pathogenesis model (Figure 3D), all the four genes should be in the same ontology with either *GREM2* or *GPR146*, but not both, except there were *PIP5K1B* and *EZR*. The former was downregulated, while the others were upregulated, which might or might not result in EOPE. Nonetheless, the opposite regulations resulted in normotension without FGR. As implied by GSAI annotation (Table S3), the downregulations of the four genes lead to: (1) less mRNA processing and export and adipogenesis by ALKBH5; (2) less activation of RAC1 by SWAP70, which regulates actin cytoskeleton dynamics; (3) less dephosphorylation of MAP kinase by DUSP1, involved in cell proliferation, differentiation, and response to stress; and (4) less transcription of genes by KLF4, which play roles in specific processes involved in cell cycle regulation, apoptosis, and epithelial mesenchymal transition. Meanwhile, the downregulations of *GREM2* and *GPR146* respectively lead to: (1) less antagonistic effect on bone morphogenetic protein involved in organogenesis; and (2) less G-protein coupled receptor with unknown exact function. Respectively, the up- and downregulations of *PIP5K1B* and *EZR* lead to: (1) more phosphatidylinositol 4,5-bisphosphate, a critical component of the inner cell membrane involved in signal transduction and cytoskeleton organization; and (2) less linkage of link the actin cytoskeleton to the plasma membrane, involved in cell adhesion, migration, and signal transduction.

## Discussion

### Finding summary

Our pathogenesis models connected both endometrial maturation and HELLP syndrome with LOPE and EOPE. The former was connected with chorioamnionitis, while the latter was connected with placentation. Several biological processes were indicated by the hierarchical learning representation of the models.

In the pathogenesis models of LOPE, the hierarchical learning representation indicated three biological processes. The first process indicated more mitochondrial ribosome biogenesis and cellular respiration in third-trimester placenta, which were more similar to endometrium during proliferative phase than mid-secretory phase. The second process indicated less placental cell growth, differentiation, and survival, and more response to hypoxia, which were the opposite to chorioamnionitis. However, it was similarly less T-cell overactive response or protein degradation via ubiquitination. The third process indicated downregulation of vascular tone in third-trimester placenta, similar to HELLP syndrome.

Meanwhile, the hierarchical learning representation of EOPE pathogenesis models indicated the roles of four biological processes in protecting against this condition. The first process indicated less placental intracellular lipoprotein, less cellular resistance to mechanical stress, and less immune suppression via regulatory T cells during third trimester, similar to endometrium during proliferative phase compared to late-secretory phase. Except, it indicated less lipoprotein secretion via SNAP23. The second process was similar to the normal biological process in first-trimester placenta compared to term placenta. This biological process indicated more placental intracellular magnesium as a critical cofactor for many enzymes, resulting in pyruvate and glutamate. They might be either from: (1) the products of misfolded-protein degradation, or (2) metallothioneins (high cysteine residues). The latter also indicated more placental cell development and transport, and less vasodilation effect of nitric oxide. Independently from such indications, the second process also indicated less T-cell activation and differentiation but more migration and adhesion of monocytes, macrophages, and T cells during third trimester. The third process indicated less placental cell growth, development, and metabolism, less remodeling of extracellular matrix. Such indication was similar to the normal biological process in second-trimester placenta compared to term placenta. The difference was less potassium-dependent exchange between sodium and calcium to maintain their concentrations in placental cells during third trimester. The fourth process indicated less mRNA processing and export, less dephosphorylation of MAP kinase, less actin cytoskeleton dynamics, and less epithelial-to-mesenchymal transition. The cytoskeleton dynamics might be related to either more cytoskeleton organization or less placental cell adhesion and migration, but not both. Except, there was less antagonistic effect to organogenesis and less G-protein coupled receptor. These biological processes were indicated to be necessary in protecting against this condition, but the protection loss was not sufficient to lead to EOPE, except there were additional factors.

### Comparison to previous studies

Almost all the biological processes (*n*=6/7, 86%) in this study included at least one gene that was supported by previous human studies [32–56]. A biological process, that included MRPL34 and DYNLL1, was a novel finding for LOPE, which was similar to endometrium during proliferative phase compared to mid-secretory phase. For this subtype, the other genes were also novel findings (i.e., *ANKRD13A*, *SLA*, and *FAM43A*). Meanwhile, we also found novel genes for EOPE across all the biological processes (i.e., *SNAP23*, *PPL*, *LRRC32*, *GPT2*, *UBE2H*, *NIPAL3*, *NIN*, *KIAA0232*, *MT1F*, *DKK3*, *SLC24A3*, *SWAP70*, *GREM2*, *GPR146*, *PIP5K1B*, and *EZR*). A gene for each subtype was frequently studied in the context of PE as the dependent variable, i.e., IGF1 [36–55] and PAPPA2 [57–68] respectively for LOPE and EOPE.

For LOPE, our results indicated less placental cell growth, differentiation, and survival via IGF1. Previous studies provided a sequence of evidences of the impact of *IGF1* downregulation on placenta structure and function in LOPE. First, lower IGF1 was consistently found in placental samples [44], villous and extravillous trophoblasts [42], and maternal and cord blood [40] from women with LOPE compared to those from healthy pregnant women. Second, silenced *IGF1* in CD-1 mice resulted in: (1) elevated systolic and diastolic blood pressure and urine protein; (2) reduced placental vascular density and branches and fetal weight; and (3) placental structure derangement [54]. Third, reduced IGF1 in trophoblastic cell lines subsequently led to reduced viability, reduced capabilities in cell proliferation, migration, and invasion, and enhanced apoptosis, including cell lines representing extravillous trophoblasts [47, 50, 52–54]. The effect of IGF1 on trophoblasts were mediated by reducing ZEB1 via Erk signaling pathway [54]. *ZEB1* was also downregulated in placental samples from women with LOPE compared to healthy pregnant women, and reduced ZEB1 simultaneously led to downreguled hsa-miR-183 and reduced cell viability, migration, and invasion [54].

Previous studies identified multiple antecedents of IGF1 reduction in placenta. First, reduced IGF1 was directly associated with hypermethylation at a CpG site of IGF1 promoter, preceded by elevated DNMT1, particularly under hypoxia, in which DNMT1 expression was also higher in placental samples of LOPE than those of healthy pregnant women [37]. Second, hypoxia and reoxygenation also upregulated exosomal hsa-miR-486-5p which lead to reduced IGF1 and cell viability, proliferation, migration, and invasion in trophoblastic cell lines [50]. Third, another miRNA, i.e., hsa-miR-206, was consistently upregulated in maternal blood, myometrial, and placental samples [38], and maternal blood and decidual samples [46] from women groups with either exclusively or predominantly LOPE compared to those from normotensive pregnant women during third trimester. The aforementioned upregulation of hsa-miR-206 was accompanied by downregulated *IGF1* in placental samples [38] and reduced IGF1 in maternal blood [46]. Upregulated hsa-miR-206 was also accompanied by downregulated *MALAT1* in placental samples from women with PE compared to those from healthy pregnant women, in which MALAT1 binded and negatively regulated hsa-miR-206 expression and it subsequently reduced IGF1 via PI3K/Akt signaling pathway [53].

For EOPE, our results indicated less cleaving of IGFBP5, which in turn, reducing of released IGFs involved in less cell growth, development, and metabolism, and less remodeling of extracellular matrix via PAPPA2. As a biomarker, PAPPA2 in maternal blood demonstrated higher sensitivities for EOPE (∼60% [58] and ∼40% [62]) than those for LOPE (∼4% [58] and ∼19% [64]) at ∼95% specificity. PAPPA2 in maternal blood and uterine artery pulsatility index were also observed higher in Andean pregnant women living at higher altitude or with EOPE but not significantly in those with LOPE, suggesting an adaptation to hypoxic conditions [61]. Similarly, at low altitude, higher PAPPA2 level in maternal blood was observed in non-Andean pregnant women with EOPE but not LOPE during second- and third-trimester pregnancies [68]. A previous study observed an upregulation of *PAPPA2* along with *INHBA* and other 8 genes and 2 downregulated genes in syncitiotrophoblast extracellular vesicles from EOPE with FGR [57]. In placental samples from pregnant women with EOPE compared to those with normotension, higher PAPPA2 was identified in syncitiotrophoblasts and extravillous trophoblasts, which respectively led to following effects in cell lines representing them [66] : (1) reduced cell migration, invasion, and junctions; and (2) reduced cell proliferation.

Previous studies identified multiple antecedents of elevated PAPPA2 in placenta. First, overexpressed TGFβ1 led to increased PAPPA2 via SMAD3 and reduced cell migration in extravillous trophoblasts [60]. Second, *METTL14* and *circPAPPA2* were upregulated in placental samples of PE [63]. This event was accompanied with higher N6-methyladenosine (m6A) immunoprecipitation in coding DNA sequence and 3’ untranslated region of *circPAPPA2*, where METTL14 led to increased m6A and upregulated *circPAPPA2*. The latter protein led to increased invasive capability of extravillous trophoblasts [63]. Third, either inhibiting PDE5 or antagonizing EDNRA led to increased PAPPA2, INHBA, and sFLT-1 with a higher increment in placental samples from pregnant women with EOPE [65].

However, our pathogenesis models for EOPE only indicated necessary biological processes in protecting against this condition, but additional factors might be needed for leading to EOPE. In other words, the identified biomarker characteristics are probably shared with other conditions. PAPPA2 in placental samples were higher in pregnant women with LOPE [66], although PAPPA2 in maternal blood was less accurate in predicting LOPE than EOPE [58, 64]. PAPPA2 in maternal blood was also higher in pregnant women with gestational hypertension but not statistically different to that with PE [67]. Furthermore, upregulated *PAPPA2* and *FLT1* and downregulated *IGFBP5* were identified in trophoblasts of third-trimester placental samples from pregnant women contracting SARS-CoV-2 [59], a condition called PE-like syndrome [69]. Severe COVID-19 could be predicted by sFLT-1 in maternal blood [70, 71], a well-known predictor of EOPE [72]. However, PAPPA2 or FLT1 were not included in the biomarkers that predicted PE but not COVID-19, and these blood biomarkers represented maternal-fetal interface tissues [73]. Therefore, PAPPA2 is likely a common effect among pregnant women with EOPE and LOPE, gestational hypertension, and COVID-19, in which PAPPA2 reduction protected against the hypertensive property of those conditions.

### Research implication

Our pathogenesis models gained additional insights on the different roles of endometrial maturation in placentation between LOPE and EOPE. Previously, overlapped DEGs were identified for representing endometrial maturation, PE, and decidual natural killer (NK) cells [16], including *IGFBP1* and *EPAS1* (a gene that codes HIF2α). Wet lab experiments in another study demonstrated three findings confirming the overlapped DEGs [74]. First, the addition of HIF2α increased extracellular vesicle (EV) secretion by human endometrial stromal cells (HESCs). Second, the addition of EVs increased decidualization markers of HESCs, including IGFBP1. Third, the addition of EVs increased the number of extravillous trophoblasts in which *IGF2* was upregulated. In addition to IGFBP1 and other proteins, HIF2α-induced stromal EVs also carried TGFβ1 [74, 75] which led to increased PAPPA2 and reduced extravillous trophoblast migration [60]. Therefore, future experiments on endometrial EVs may reveal whether MRPL34, DYNLL1, or SNAP3 in endometrium affects gene regulation of those proteins in placenta.

Opposite gene regulations between chorioamnionitis and LOPE were also identified by our pathogenesis models. Chorioamnionitis frequently causes preterm delivery from 10.7% (*n*=53/497) to 94.4% (*n*=17/18) among pregnant women who delivered at 33-36- and 21-24-weeks’ gestation [76]. Meanwhile, PE incidence was >1 per 1000 ongoing pregnancies approximately starting from 34 weeks’ gestation [13]. However, no placental samples under chorioamnionitis were found from pregnant women with PE when we identified the gene sets of interest in this study [27]. Lacking of co-existence between chorioamnionitis and LOPE is required to whether competing risks between both conditions are merely due to clinical decision on pregnancy termination or pathophysiological due to opposite gene regulations.

For HELLP syndrome, our pathogenesis models could not differ this condition from either LOPE or EOPE. Most of HELLP syndrome cases reported hypertension which always present in PE, leaving 10-20% without hypertension [77]. Alternatively, HELLP syndrome shared similar phenotype with atypical hemolytic uremic syndrome, a condition due to complement gene mutations [78]. Similar genetic factors of mothers may lead to common effect on complement system and placenta between HELLP syndrome and PE. Investigation on these genetic factors and biomarkers in non-placental organs may help to differ PE from HELLP syndrome.

### Strength and limitations

DI-VNN method allows systematic identification of which genes contribute either LOPE or EOPE. Contribution of a gene was not only evaluated according to its expression value but also its correlation to other genes. These values were fitted to a specific PE subtype, enabling validation of the pathogenesis models using new data. Using these models, this study could differ LOPE from EOPE in the antecedents of placental dysfunction and identified the biological targets, i.e., *IGF1* and *PAPPA2*. Our models also identify other specific genes as specific context of the potential mechanism preceding placental dysfunction, that distinguishes EOPE and LOPE.

However, we are aware of several limitations in this study. First, mRNA expression findings in this study were likely the effect instead of cause of the antecedents. As we discussed, IGF1 and PAPPA2 play important roles in trophoblast migration and invasion, but it remains unclear what drives impaired regulation of these genes. Yet, our pathogenesis models also identified novel genes in LOPE and EOPE related to IGF1 and PAPPA2, which may pave the way to investigate possible driving mechanism. Second, we could not validate the pathogenesis models for other PIH and PDD subtypes using the gene sets, which we identified previously [27], due to data unavailability. The validated pathogenesis models for LOPE and EOPE might indicate the common genes with those for gestational hypertension and isolated FGR. Nevertheless, we could differ LOPE from EOPE, which is important to gain insights on LOPE to expand early intervention strategies for PE. Third, findings related to endometrial maturation could not be fully justified using our data, but we identified MRPL34, DYNLL1, and SNAP3 in endometrium which can be investigated further in wet lab experiments for their roles in placentation, e.g., evaluating their effects on stromal EVs and trophoblasts differentiation.

### Conclusions

We have developed and validated pathogenesis models of LOPE and EOPE. The pathogenesis models of LOPE revealed: (1) increased mitochondrial ribosome biogenesis and cellular respiration akin to proliferative endometrium; (2) reduced placental cell growth and heightened hypoxia response as opposed to chorioamnionitis; and (3) decreased vascular tone akin to HELLP syndrome. Meanwhile, the pathogenesis models of EOPE revealed protective roles against this condition via: (1) decreased placental lipoprotein and immune suppression akin to proliferative endometrium; (2) increased intracellular magnesium for enzyme activity and reduced placental cell growth and matrix remodeling akin to placentation; and (3) less mRNA processing and cytoskeleton dynamics akin to HELLP syndrome. However, more factors were required for EOPE in addition to the loss of these protective mechanisms.

## Acknowledgements

This study was funded by: (1) the Postdoctoral Accompanies Research Project from the National Science and Technology Council (NSTC) of Taiwan (grant nos.: NSTC111-2811-E-038-003-MY2 and NSTC113-2811-E-A49A-003) to HS; and (2) the National Science and Technology Council in Taiwan (grant no. NSTC113-2221-E-A49-193-MY3), the Ministry of Science and Technology (MOST) of Taiwan (grant nos.: MOST110-2628-E-038-001 and MOST111-2628-E-038-001-MY2), the University System of Taipei Joint Research Program (grant no.: USTP-NTOU-TMU-112-04), and the Higher Education Sprout Project from the Ministry of Education (MOE) of Taiwan (grant no.: DP2-111-21121-01-A-05 and DP2-TMU-112-A-13) to ECYS. These funding bodies had no role in the study design; in the collection, analysis, and interpretation of data; in the writing of the report; or in the decision to submit the article for publication.

## Declaration of Interests

HS has signed an expert agreement with Atheneum. The other authors declare that they have no competing interests.

## Data Statement

Data are available publicly available in GEO and ArrayExpress, as described in our previous work [27] (https://github.com/herdiantrisufriyana/pec/blob/master/supplementary_material.pdf). We also publicly shared the source codes for data analysis (https://github.com/herdiantrisufriyana/pec).

## Authors Contributions: CRediT

**HS:** Conceptualization, Methodology, Investigation, Data curation, Validation, Formal analysis, Writing – original draft, Visualization, Funding acquisition. **YWW:** Conceptualization, Methodology, Writing – review & editing, Supervision. **ECYS:** Conceptualization, Methodology, Resources, Writing – review & editing, Supervision, Funding acquisition. All authors have read and approved the manuscript and agreed to be accountable for all aspects of the work in ensuring that questions related to the accuracy or integrity of any part of the work are appropriately investigated and resolved.

## Notes

### Author Declarations

Data are available publicly available in GEO and ArrayExpress, as described in our previous work (https://github.com/herdiantrisufriyana/pec/blob/master/supplementary_material.pdf).

