## Supplemental Materials for "Hierarchical representation learning of preeclampsia interactome connecting endometrial maturation, placentation, chorioamnionitis, and HELLP syndrome"

Table S1. Predictors in the pathogenesis models.

| Subtype | | Representation of | | HGNC symbols |
| --- | --- | --- | --- | --- |
| PE | Early-onset, severe | Endometrial maturation | Early-secretory phase | FBLIM1, PALLD, INPP5F, PPT1, OAZ1, HEXB, PDGFC, OLFML2B, HMMR, SPON2, TOP2B, DCP2, SCARB1, UBE2Q2, HTRA1, LSM6, MOB3C, ILF2, CKS2, SLC23A2, XPO4, TEX2, GOLM1, TBX3, SLC29A3, ENG, SETX, KDM1A, TES, GNG7, QPCT, PM20D2, AMOTL2, FPGS, FSCN1, SLC7A2, TYRO3, SPON1, DDHD1, TSPAN33, MGAT5, CREB1, AGO1, CNPY4, CIB2, CXXC1, SYK, SPG21, NRSN2, PSAP, LRRC42, RABEP1, DDR2, RNF24, HJURP, CBX5, MID1, YWHAG, CDK14, TMEM30A, CYB561A3, KLHL3, PROCR, CNN3, IRF8, PIP5KL1, CCND3, SLCO2A1, PRRX1, MIIP, PFKM, SHANK2, HMGB3, ZNF462, GPX7, GPX3, COQ2, CARD16, SH3PXD2A, DROSHA, DNAJB6, CD47, STX6, LHFPL2, NRCAM, VCP, FRZB, CYFIP2, SESN3, PMAIP1, MELK, EZH2, GNG4, TNFRSF10D, VEGFA, FBN2, RAD51, MAP4K4, TRIP13, INTS7, DNMT3A, EFNB1, TRIB2, ALDH1B1, MICU3, LATS2, BASP1, FANCE, ODF2, RPL15, TRPC6, GRAMD1A, SLC26A6, TRIM25, SPRED1, MX1, KLHL42, OSMR, LMAN2L, LAMC2, SH2B3, SLC8A1, IL33, PKM, OVGP1, IGSF3, DLGAP4, SYDE1, DPYSL3, CDC42EP1, THOP1, TMEM216, ARHGAP28, TMEM106B, SH3BP5, PPP2R3A, MXRA5, RPS6KA6, APLN, LAMA4, MBTPS2, NCEH1, HNRNPA0, DIO2, RUSC2, LYN, PGRMC1, DONSON, CTSV, ETNK2, PANX1, PAWR, SLC6A8, GNG11, SUMO3, MTPN, CEBPG, ROBO4, RAB5B, FEM1B, CAPRIN1, ZMYND11, PUS7, TXNRD1, NUDT5, PAK2, ACTR2, PLBD1, RASSF8, GNB1, ZEB2, PTPRD, KIFC3, GRINA, MX2, TAF9B, TNS3, CORO1A, PLCB2, LEPROTL1, ATXN10, CDO1, RGS10, RCBTB2, HMOX1, MED13L, OPTN, LRRC32, ITGA5, PKD2, PTTG1IP, NUFIP2, IFIT1, TNFAIP8, NRIP1, PROM1, ZNF512B, UNC119, SIPA1L1, MUC15, MAD2L2, RNF20, RIN2, CDC42BPA, GPER1, PHTF2, RIOK1, RSRC1, SIK3, SUV39H1, RAB6B, RAB8B, CTBS, ARHGDIB, PARM1, FBLN7, SHMT2, RANBP6, PAPPA, HDAC1, SSR1, OAS3, CLOCK, RNF114, CDC42BPB, FOLR1, MLLT1, CPEB4, ARID3A, ZNF84, GRHL1, ST6GALNAC1, GLA, PJA1, LRCH1, RAPH1, AKAP13, SAMD8, NFKB1, ATP1A1, MYO5A, HPCAL1, MME, HEXIM1, SNX18, MAOB, MEF2A, DCXR, SERPINB9, ABAT, CITED2, LRRC1, SLC44A3, KLF4, CEBPD, BRCA1, FOSL2, TRA2B, SLIT3, CLIC5, GCLM, OPRK1, PLPP2, FSTL3, MAP3K8, CALM1, SPAG4, TIMP3, TRIML2, EXOC6, SLC39A8, FAM210A, TMEM141, HDHD3, PRKAB2, GPR146, EPHB3, MIOS, MMD, TM9SF3, ARHGAP26, PLIN2, RERG, ZFR, ZDHHC9, MYO9A, APOL4, ERP44, NEK11, RNASE1, NFIL3, DKK3, TRIP4, FAM120AOS, CLMN, FAM107B, WWC1, RBM47, NPC1, BHLHE40, DUSP1, S100A6, PTPN3, CDS1, SGMS2, TLCD1, RNF19A, EPS15, WDR11, PHKA2, TACC1, ABCA1, RHOBTB3, CRYAB, EDN1, EFR3A, LURAP1L, ARTN, GPD1L, PAPSS1, CDYL2, TMCC3, SH3RF1, SBF2, C9orf72, CNOT8, SORT1, OCEL1, WDFY2, GADD45G, SLC17A5, WDFY3, PPA1, PLCB1, TSPAN12, LRIG3, G3BP2, DISP1, BCAT2, KLF3, FBXO8, FLNB, PLPP3, NFIA, EZR, ZNF217, ANGPT1, KIAA0232, ERCC6, GULP1, ANXA1, JADE1, DUSP4, ZNF143, ERRFI1, CITED4, SPTY2D1, TSPAN13, NUP153, PLOD2, VSNL1, TMEM154, GSN, PTPRJ, SEMA3G, PPP2R1B, SACM1L, FRMD4A, LEP, TANK, PRKCH, ARHGAP5, BTD, CANT1, AMOT, CYP2J2, NHLRC2, NECTIN4, HSPA2, UBALD2, IPPK, EAF1, TPD52L1, PLEKHA7, AVPI1, DPYD, P4HA1, TRIM52, PDZD2, GDA, ADCY7, CDKN1B, PGBD2, PTBP3, FZD1, MATN2, APOL2, ARRDC3, LRRC41, COL1A2, PTPN12, ELL2, SLCO4A1, FLVCR2, SAV1, MXI1, TMEM268, SLC24A3, GPT2, OSBPL11, SNAPC1, CYB5R1, GGCX, PPP1CB, SLC31A2, RASEF, IQGAP2, PPP3CA, ACVRL1, CX3CR1, PREP, ARL6IP5, SRPK2, HMGCR, CRISPLD2, MRI1, TLK1, PPL, MAFF, PIP5K1B, DSP, GOT1, SLC2A1, SWAP70, CCSAP, SLC35B3, NEK7, ERLEC1, KRT19, ESRP1, TGFBR3, SCAPER, EPS8L2, DDIT4, TIGD2, RALGPS1, HILPDA, BCAR3, ANKRD37, MXD1, ADK, ICA1, GOLGA5, TLR4, TSPAN31, PIK3CB, ANKRD13A, HEY1, PRRG4, DLX5, ZBTB20, MT1F, SNAP23, WDSUB1, NT5E, AMD1, VEGFC, C1orf21 |
|  |  |  | Late-secretory phase | ABCA1, ANKRD13A, ZFAND5, PANX1, RASEF, LAPTM5, RNF11, ACKR2, TBXAS1, ETS2, ARRDC3, BHLHE40, MTMR6, ARHGEF12, STRN3, ELF4, RAPH1, ENG, EPS8L2, RGS16, GRHL1, MBD2, OXSR1, CTSA, NCEH1, VEGFA, AKAP13, TMEM154, LDLR, JUNB, YPEL2, CDYL2, TES, MAFF, CEBPD, GALE, SMIM3, SH3BP5, LYN, CSF2RB, TSC22D1, GULP1, CDKN1B, GADD45G, CORO2A, GPX3, NEK7, PRRG4, MINPP1, MXD1, SLC1A1, FAM43A, ZNF217, CHIC2, PPP2R5A, SLC2A1, PIK3CG, ZNF281, PIK3AP1, SLC38A6, TMEM176B, SNAPC1, SPTY2D1, HILPDA, IFIT1, STAB1, NR4A3, ZFYVE1, TIMP3, MAP3K8, CORO1A, SYNJ2, SLC38A1, RFK, PRKCH, RNF19A, ERP44, PPL, PGBD2, LATS2, HTRA1, LRRC8A, PAPPA, TREM1, KPNA1, TYROBP, MXRA5, NUCB2, LCP1, GBE1, PGK1, CPEB4, LURAP1L, EAF1, DUSP4, FBXO9, TSPAN12, RBM47, OPTN, ANKRD37, VDR, FAM210A, SLC25A4, MXI1, TMEM216, DUSP1, SLC7A2, TEX2, PIK3CB, PLIN2, LPL, SAMD8, ITGA5, TSPAN13, KIAA0513, IGSF3, ST6GAL1, SPAG4, SH2B3, ANXA1, PPA1, HEY1, TGFBR1, POLR2H, TANK, ILF2, STAT4, LYPD3, WDSUB1, KDM1A, PREP, CX3CR1, TPP1, RNASE1, GNG4, SGMS2, THRA, TMEM30A, DDIT4, SEMA4B, CASD1, PTPN12, FKBP4, DOCK8, KLF4, THOP1, UBR1, EPS8L1, BCAR3, SPI1, TP53BP2, SWAP70, FLI1, C9orf72, TSPAN14, SUMO3, PDZD2, CTBS, TMCC3, EDN1, ZNF143, FNDC3A, SH3PXD2A, TRAK2, ALDH1B1, DLX5, MYO5A, GRAMD1A, SLCO2B1, CIB2, RHOBTB3, DNAJB9, COL1A2, ARHGDIB, GLA, SARAF, NFIL3, HDAC1, MRI1, CYFIP2, MED13L, WDFY3, SMPDL3A, DCXR, EPS15, MEF2A, EFHD1, QPCT, HNMT, AMPD3, CRISPLD2, KIF17, CDK9, SLC9A9, C3AR1, FBXO8, LSM6, GNG7, GPER1, USP3, AMD1, PTBP3, COL17A1, GLE1, FOSL2, ELL2, LRRC32, ZNF512B, SLCO4A1, COQ2, COL6A1, PHKA2, CRYAB, CLIC5, HPGDS, SLC31A2, LIN7A, LY86, UBALD2, TNFAIP8, CTSF, COPZ2, TOP2B, CTSV, RNF24, SLC17A5, ARHGAP28, LRRC41, RCBTB1, PROM1, SERPINB9, SLC16A3, MAOB, SPON1, ASTE1, NPC1, DONSON, NARF, TNFAIP8L1, FAM172A, BRCA1, CHEK1, SBF2, ELMO1, TSPAN3, GRINA, KIAA0232, ASAP2, TMEM106B, ODF2, KLF3, XPO4, CDO1, LPXN, SULF2, MYO1F, DNMT3A, CALM1, MTPN, HLF, SH3BGRL, CLIC3, HSPA2, WFS1, NRIP1, SESN3, PIK3R1, TBC1D5, FGF13, RIN2, TYRO3, RBM26, TRIM25, HEXB, ARHGAP24, MAP4K4, CDK20, CYBB, OVGP1, TAF9B, OSMR, SMARCA2, DPYD, SACM1L, PLOD2, SNAP23, DNASE2, STK17A, CASP4, IGSF8, SUV39H1, CNPY4, TTLL5, ERO1A, OAS3, GOSR1, HMGB3, TRIM24, EDEM1, PPM1M, RAB8B, ST6GALNAC1, CCND1, MAD2L2, SLC39A8, FZD1, BAD, MOB3C, CLCN5, C4orf3, RDX, CSF3R, PCYOX1, PUS7, DDX17, FBN2, PPT1, PJA1, PEBP1, IL33, TKFC, CBX5, SRPK2, FBLN7, PALLD, CDC42BPA, CNN3, FAM107B, DDR2, DEGS1, GEMIN5, PODXL, FBLIM1, AMOTL2, LHFPL2, MAML2, PDGFC, DKK3, MID1, NIPAL3, PCCB, ATXN10, HJURP, SHANK2, TOR1B, PKM, BCAT2, CCDC113, UNC119, MME, MELK, RAD51, HMMR, MICU3, COX8A, INPP5F, PLBD1, MX1, CPD, ZNF462, GREM2, RNF20, TPD52L1, MTA1, PTPRJ, CKS2, TRIP13, PIP5K1B, LMAN2L, PLCB1, IGF1, FRMD4A, AZIN1, ZFP3, FOLR1, UBE2K, INTS7, CITED2, HNRNPA0, MATN2, GPX7, HMGN3, ZMYND11, PAM, HMGCR, MBNL2, EIF4A2, RPL15, CCDC115, NRCAM, PIGX, GSTK1, PIP5KL1, TRA2B, EPHB3, PFKM, ZNF84, PGRMC1, SLC23A2, STXBP6, CLOCK, TRIB2, DROSHA, CXXC1, NFIA |
|  |  | Placenta | First trimester | CX3CR1, SLIT3, CEBPD, MYO5A, SLC2A1, GNG11, DNAJC1, AKAP13, SCARB1, ZBTB20, PTPN12, IGF1, KIAA0232, CBX5, DYRK2, ETS2, MEF2A, GPER1, ZNF462, IFI44L, CYTL1, GSN, PARM1, NFKB1, RAB3B, SEMA3G, ST6GAL1, MCTP2, SYK, NFIA, TXNDC11, SMIM3, LRRC8A, JAK2, CDK14, MS4A4A, LAPTM5, JUNB, F8, RIN2, CLIC5, CARD16, PRKCE, KIF5B, CCDC59, TBXAS1, DDIT4, GNB1, EXOC6, CCND1, TMTC4, SAV1, NECTIN4, PPP2R5A, KMT2E, FZD1, CYBB, DPYD, ELMO1, SNAPC1, C3AR1, TSPAN33, HMGN3, MIDN, LTF, RIMKLA, TRIP10, CLOCK, TACC1, FLI1, MBTPS2, MXI1, RNASE1, MAML2, SYPL1, AMOTL2, DDR2, ACTN1, TDP1, MIIP, TANK, ARRDC3, YPEL2, GALNT18, TYROBP, DUSP1, ZNF281, LAMA4, EFHD1, EDEM1, SBF2, TEX2, THOP1, PPP3CA, TLR7, THRA, LRRC32, HSPA2, PIK3AP1, TSPAN14, ZFYVE1, MRI1, CHIC2, MLLT1, COL6A1, SLC9A9, MINPP1, DIO2, SULF2, ASTE1, PLCB1, RNF11, CMKLR1, ZNF331, PODXL, SDC4, WDSUB1, HMOX1, PDGFC, SMPD2, SLC26A6, PGRMC1, SLC6A8, GUCA2A, TLR4, BCL2L11, MXRA5, MAP3K9, BCORL1, HK1, RCBTB2, ASAP2, RALGPS1, ADCY7, TMCC3, ANGPT1, ARL6IP5, RASAL2, ARTN, RASEF, ANKDD1A, AVL9, SYDE1, MATN2, ILF2, GCLM, CTBS, GOLM1, PROM1, PM20D2, EBF3, SH2B3, ATP10D, IL33, ERRFI1, HNMT, DKK3, ELL2, GRINA, PDE5A, MAP4K4, SESN3, NMNAT2, BRAF, INSL4, NIPAL3, TRIB2, ARID3A, KDM1A, TMEM106B, VEGFA, CNNM4, WDR11, MT1F, CREBL2, PKM, CDO1, NUDT5, HEY1, PEBP1, SLC38A1, LHFPL2, NCEH1, SEMA4C, ZMYND11, PLOD2, TRPC6, FANCE, VEGFC, PMAIP1, SHANK2, PFKFB4, ACVRL1, SLC35B3, GALK1, GBP4, EDN1, NUCB2, DHX29, ADARB1, OLFML2B, CPOX, PAPSS1, GPR107, HMGCR, MIOS, ACSF2, DLX5, CCDC113, GAN, TMEM176B, FCN3, ARHGAP24, CDS1, KIFC3, XPO4, LRCH1, PCYOX1, INTS7, AZIN1, SPG21, APLN, WNK2, G3BP2, FGF13, SLCO2B1, LIMCH1, AGTR1, FBN2, SARAF, CNOT6L, TRIP13, YTHDC2, ZNF512B, TMEM141, TAPT1, SACM1L, INSIG1, RNF20, BRCA1, ERLIN2, CADM3, AMD1, GLA, PUS7, CKS2, ROBO4, AK2, PRKAB2, BTD, HMMR, PLBD1, LEPROTL1, WDFY2, FLVCR2, PHKA2, PDP2, DCXR, SLC38A6, SLC39A8, NPL, SLC11A1, CTSA, PTPRJ, C1orf21, MME, DUSP4, MXD1, KLHL3, RCAN3, APH1B, FOLR1, DUS2, SLC44A3, TMEM43, PIK3CB, ABLIM3, GRHL1, DAB2, LDLR, SIPA1L1, SLC24A3, MAT2A, MKNK1, SLC16A3, GRHL3, EPS8L1, HGF, TP53BP2, LSM1, MOB3C, NHLRC2, CDYL2, KIF2A, MAN1A2, PAPPA, NPEPPS, INPP5F, NEK7, TLCD1, RASSF8, TMEM268, CSNK2A2, CCSAP, KIF17, ZNF75A, KLHL42, PKD2, NIN, PPP2R1B, HSPA9, GGCX, CYP19A1, GPT2, SH3RF1, EZH2, TKFC, LURAP1L, CHEK1, SPON1, SCMH1, PLAG1, STX6, EPB41L5, DONSON, IDE, PDZD2, GEMIN5, PRKCH, RAD51, MAFF, GPD1L, TOP2B, PREP, ARHGAP26, FNDC3A, STAT5B, CLTC, GADD45G, ODF2, VCP, RHOBTB3, TMEM154, DOCK8, MELK, SMARCA2, TTI1, MTMR4, ETNK2, ARHGAP28, ESRP1, HJURP, UBE2H |
|  |  |  | Second trimester | GADD45G, KIF2A, STAT5B, HEY1, TMTC4, SLC35F2, ENG, C1orf21, LIMCH1, CLIC5, CLMN, TAPT1, APLN, SCARB1, HMGN3, GPER1, PAM, EPS8L1, MAFF, VSNL1, PAPPA2, PRKCH, NECTIN4, SLC7A2, SLC24A3, RASAL2, CCSAP, NEK7, PLOD2, PAPPA, CDK19, ABLIM3, MTMR4, HJURP, ARHGAP26, SYDE1, DUSP4, ACSF2, MAN1A2, EFHD1, PLCXD2, HMGCR, GOLM1, RAB3B |
|  |  | Chorioamnionitis | Second trimester | PROCR, CCDC113, FSTL3, RASEF, CNNM4, NEK11, GPT2, LIMCH1, LPL, LEP, CX3CR1, SH3PXD2A, INHA, DLX5, LRRC1, SLCO2A1, KRT19, QPCT, PIK3AP1, ENG, SYDE1, VDR, FCN3, ERRFI1 |
|  |  |  | Third trimester | C4orf3, EAF1, LEP, OLFML2B, SUV39H1, TSPAN33, INHA, SEMA4C, PIK3R1, COL17A1, FKBP8, RNASE1, ZMYND8, CD4, TBXAS1, MXI1, SERPINB9, NFIA, FSTL3, HILPDA, ANKRD37, GREM2, NRIP1, MID1, UBB, CDO1, ITGA5, SIPA1L1, SYK, FAM20B, PLCB2, TMEM45A, DIO2, ARRDC3, RFK, P4HA1, SEC23A, RNF24, PMEL, HPCAL1, ERRFI1, COL6A1, COL1A2, RAPH1, DNAJC1, SPON1, GBE1, KCTD12, BTD, GPT2, SLC11A2, TREM1, CEBPA, PDGFC, AMD1, MFAP5, CXXC1, BHLHE40, VCP, TRIM24, RASSF8, BCL2L11, DISP1, VDR, PPA1, KMT2E, ZC2HC1A, SLCO2A1, MS4A4A, CCDC127, SYNJ2, MAP3K8, ERO1A, SLC2A1, MGAT5, GALK1, DCP2, EBI3, PROM1, GDPD3, TMEM107, MAP4K4, RGS10, INPP5B, TSC22D1, GNB1, DPYSL3, PTBP3, TNFAIP8L1, AVL9, DSP, GSTK1, IPPK, BRCA1, KIAA0232, MKNK1, TMX1, APOL4, KIF2A, PAM, CYB5R1, PUS7, CNNM4, LPL, DOCK8, TNFAIP8, PFKM, TANK, HMMR, CTSZ, PAWR, CASP4, CHEK1, SDC4, NUFIP2, FBN2, ENG, HMGCR, INTS7, UBE2H, KLF4, SDF2L1, GPR146, CORO1A, F8, FSCN1, VEGFC, ARTN, FBLIM1, RAP1GAP, FAM107B, EDN1, ERP44, OSBPL11, UBR1, SLC35F2, GADD45G, PODXL, ALKBH5, SLIT3, GFOD2, AMOTL2, COQ2, PPP2R5A, TMEM43, ZNF175, CCND1, QPCT, PJA1, TACC1, TSPAN3, OCEL1, LAPTM5, CALM1, YPEL2, RIOK1, ELMO1, ABCA1, MAT2A, KLHL8, MATN2, LRRC32, RANBP2, SMPD2, GSN, NECTIN4, IFIT1, NARF, SHANK2, STXBP6, IFI44L, SHMT2, SLC11A1, ICA1, GBP4, CADM3, LTF, ARID3A, FOXJ2, PTPRD, TSPAN13, RBM22, TMEM141, SYNGR1, SPRED1, MX2, CLCN5, FBLN7, LURAP1L, PIK3CG, PALLD, SLCO4A1, ST6GAL1, PEBP1, ARHGAP28, ALDH1B1, STAT5B, BCAR3, LDLR, PAK2, SMPDL3A, NEK11, GPX7, TYROBP, ARMCX1, TBC1D9, TTLL5, CYTL1, PIK3CB, PKD2, TES, RBM47, SDF2, TDP1, EPS8L2, SLC30A1, CKS2, ILVBL, ASTE1, CCSAP, SIGLEC6, APLN, KLHL42, ILF2, OVGP1, PGBD2, FGF13, MTMR4, AMOT, ROBO4, SNX18, CNN3, SPAG4, WDFY2, SBF2, PRRG4, INSIG1, SARAF, HJURP, STAB1, GPD1L, SLC24A3, RAD51, KDM1A, PLAG1, LRRC42, LMAN2L, JUNB, SLC6A8, FLI1, ST6GALNAC1, GOT1, RBBP4, CDS1, LEPROTL1, PRRX1, MB21D2, PMAIP1, CHMP2B, ANXA1, GDA, PLPP3, SPTY2D1, PDZD2, ASAH1, COPZ2, TRIP13, ARHGDIB, TMEM30A, SMARCA2, MELK, IGSF3, ACVRL1, ADCY7, C9orf72, SSR1, CBX5, GPER1, SGMS2, ZBED5, SH3BGRL, CEBPG, FAP, TLCD1, SLC17A5, EPB41L5, FAM120AOS, ATXN10, ANXA4, SEC22C, KIF17, ZNF555, TNFRSF10D, CDC42BPA, RAB6B, ATAD1, BCL2, PIGX, STK24, DNMT3A, SEMA3G, SLC38A6, S100A6, RNF11, CX3CR1, ACTR2, ZEB2, VWCE, ACTN1, ZNF79, STAT4, RNF20, CLTC, SFXN3, GOLGA5, SLC39A8, PLIN2, ERLIN2, MRPS30, GNG11, NPL, RASEF, NPC1, SH3BP5, C3AR1, EZH2, XPO4, SCMH1, WFS1, MYO5A, ACVR2B, ZFAND5, GAN, LY86, PAPPA2, AKAP13, LRRC1, MIDN, FANCE, HDHD3, PRKCH, GNG7, ZNF143, SLC9A9, CALM2, CDK19, NRSN2, SLC7A2, EPS15, GRINA, CD28, LIMCH1, HPGDS, SPI1, CSF3R, GRAMD1A, CREB1, SLC28A1, MBD2, FOXJ3, EXOC6, PRKCD, RERG, CLIC3, VSNL1, CYBB, YTHDC2, MYO1F, HGF, MOB3A, HMGB3, RCBTB2, DLX5, CORO2A, WDR11, GRHL3, LYN, HSPA9, RPL15, MCTP2, TLR7, CMKLR1, IDE, DUSP1, SLC16A3, TRAK2, IGF1, SH3PXD2A, ANKRD13A, CCDC115, GRHL1, BAD, SAR1A, PIK3AP1, HTRA1, MAFF, RUSC2, MED13L, PROCR, RIMKLA, HNMT, MAML2, CARD16, PPP1R12C, HEY1, SLCO2B1, GUCA2A, EZR, CCDC113, EFHD1, MXD1, EFNB1, GALE, UBE2Q2, BCAT2, SYDE1, EPS8L1, KRT19, PPL, SCARB1, TRIP10, LRRC8A, ACKR2, CSNK2A2, MAP3K9, P2RY2 |
|  |  | HELLP syndrome | Third trimester | GPR146, SUV39H1, PIK3CB, CORO2A, BASP1, DLX5, SFXN3, SLC6A8, ZMYND8, SEMA4C, LRRC8A, CNNM4, HEY1, SEC23A, MIDN, EAF1, ERRFI1, PFKFB4, SLC2A1, VWCE, ANKRD37, ITGA5, GALK1, FANCE, MME, PPL, SWAP70, CYP2J2, MOB3A, SEMA4B, FBLIM1, ALKBH5, EFNB1, TSC22D1, KRT19, GUCA2A, C4orf3, SPAG4, SLC16A3, NECTIN4, TPP1, BCAT2, EPS8L2, GREM2, KLF4, DUSP1, DNAJC1, PIK3R1, SMIM3, GLA, NPEPPS, EPHB3, CSNK2A2, SIPA1L1, TOR1B, CLIC3, WFS1, CEBPA, PGRMC1, PLIN2, MYOF, TMEM45A, PLOD2, KLF3, EPS8L1, IGSF8, GRHL1, CXXC1, ENG, ARID3A, FLNB, TIMP3, PPP1R12C, AFMID, PIP5KL1, ELF4, FSTL3, INPP5B, BCORL1, APH1B, INHA, MAP3K9, SYDE1, PMEL, SHANK2, TREM1, IL36RN, HK1, FCN3, AMOT, QPCT, GSN, CCDC127, IPPK, ZNF331, NIM1K, GRHL3, HSPA9, GDPD3, P2RY2, ZDHHC9, MAML2, PIGX, ACKR2, WNK2, CDO1, TM9SF3, P4HA1, PREP, CYB5R1, SLC26A6, PRKAB2, MAP3K8, VEGFA, OVGP1, SLC31A2, TSPAN12, GCLM, PLCXD2, EFHD1, OCEL1, ZFAT, SCARB1, BHLHE40, FOSL2, SLC25A4, JUNB, LRRFIP2, HMOX1, KIF5B, SDC4, BAD, DPP7, WDFY2, FPGS, PRKCH, CLTC, CDK9, TMEM107, DONSON, ERO1A, NEK11, KIT, SPG21, GPT2, LDLR, HPCAL1, RPL15, TRIP10, ZC2HC1A, ABCA1, KMT2E, PHTF2, DIO2, PODXL, MBNL2, GORASP2, MXD1, STK24, CRISPLD2, NUFIP2, ZNF143, VEGFC, RSU1, IQGAP2, SUMO3, CREB1, FRZB, BCAR3, EBI3, NOD1, ASTE1, HMGB3, TXNRD1, SH3BP5, IGSF3, RASEF, MOB3C, TAF9B, PFKM, SH3PXD2A, PAWR, BTD, FKBP8, MTMR6, PIK3AP1, IGF1, PGBD2, MUC15, IDE, CCND1, MIIP, MAP4K4, PCCB, SH3RF1, FRMD4A, TLE6, CLDN16, PKD2, GGCX, MAT2A, NFIL3, ANXA1, NMNAT2, RNF20, ASAH1, CEBPD, C9orf72, GPX3, HSPA2, PRKAR1A, DYRK2, LYN, ARRDC3, MRS2, FEM1B, ERCC6, PDE5A, IL33, FBXO8, PJA1, SPRED1, PIP5K1B, APLN, GBE1, BTBD10, LEP, TSPAN3, ILVBL, CYB561A3, EXOC6, PAM, GALE, ZNF462, DDR2, STXBP6, PKM, HTRA1, LIMCH1, ARHGAP5, FAM43A, ADK, ATXN10, HPGDS, CALM1, ZFR, FAM20B, TRA2B, PAPPA2, CASD1, WDR11, RANBP6, SACM1L, CYP19A1, SLC23A2, UBE2Q2, HNMT, ZEB2, TRIM24, MXRA5, AZIN1, RBBP4, CCSAP, CD28, SPON1, PROCR, LRRC1, TLR7, UBB, KLHL42, ZNF555, CARD16, ZNF75A, AK2, NHLRC2, ZMYM4, VWA5A, PLPP3, MRPS30, FBN2, ZMYND11, CYBB, EZR |
| PE | Late-onset, severe | Endometrial maturation | Mid-secretory phase | ICA1, PLIN2, BMP2, DOCK8, GDA, NFIL3, RDX, CCDC127, CORO2A, BCAR3, SPAG4, FAM172A, ANKRD37, PKM, CLIP1, TUBB4B, AZIN1, ANKRD13A, PDE5A, IGF1, TUFT1, KRT19, INPP5F, EFNB1, SMPDL3A, PTPRD, NPR3, P2RY14, CTBS, SH3PXD2A, GRHL1, CDO1, LDLR, MYC, ASAH1, MRPL34, ZNF555, ELOVL5, APOLD1, FAM43A, TIAM1, BBS4, MCAT, DYNLL1, SCYL3, GPT2, SLCO2A1, SYDE1, HYAL2, TOR1B, PIK3R1, ERO1A, APPBP2, EDEM1, FAP, NECTIN4, DDIT4, NEDD4L, APLN, KLF3, GMFB, SLCO4A1, P4HA1, ERRFI1, TRIM68, ETS2, CEBPD, GULP1, RAPGEF3, FLNB, GPX3, DEGS1 |
|  |  |  | Late-secretory phase | MRPL34, DDIT4, ERO1A, KLF3, RDX, NFIL3, AZIN1, PKM, TUBB4B, SLCO4A1, VEGFA, BCAR3, SMPDL3A, CDO1, TUFT1, TMEM106B, CTBS, IGSF8, GULP1, GPX3, GBE1, EDEM1, SPON1, SPAG4, CCDC6, SLC25A4, PLIN2, NIPAL3, MYC, PREP, IGF1, INPP5F, FAM172A, DOCK8, MCAT, BMP2, PFKFB3, FAM43A, TOR1B, NFKBIA, ANKRD13A, CORO2A, SH3PXD2A, LDLR, BHLHE40, SLA, PIK3R1, TRIM68, ETS2, ANKRD37, GRHL1, P2RY14, TIAM1, DEGS1, ELOVL5, CLIP1, DYNLL1, CEBPD |
|  |  | Chorioamnionitis | Third trimester | CDO1, PLIN2, CCSAP, BCAR3, DOCK8, SPAG4, BHLHE40, NECTIN4, SIGLEC6, PTPRD, SFXN3, LDLR, CORO2A, CCDC6, TACC1, SELL, NEDD4L, SLCO4A1, SPON1, PFKFB3, GDA, APLN, IGF1, PIK3R1, SLC11A1, SLA, KRT19, SYDE1, ERRFI1, TUBB4B, SMPDL3A, GRHL1, PAK2, RBM22, CEBPA, ANKRD13A, GBE1, FAP, GPR146, HINT3, CCDC127, ZNF555, ICA1, CHMP2B, MYC, CA4, LEP, EFNB1, P4HA1, SH3PXD2A, ZMYND8, ASAH1, RAPGEF3, SLCO2A1, ZEB2, GPT2, ERO1A, ANKRD37 |
|  |  | HELLP syndrome | Third trimester | SFXN3, GBE1, ASAH1, KLF3, GPT2, LEP, SH3PXD2A, PKM, GPX3, FLNB, IGF1, CEBPD, ERRFI1, CCDC127, PDE5A, IGSF8, CDO1, CCSAP, SYDE1, ERO1A, SLC25A4, TOR1B, FCN3, BCAR3, NFIL3, LDLR, EFNB1, VEGFA, GRHL1, KRT19, ZEB2, GMFB, APLN, AZIN1, ZMYND8, FAM43A, NHLRC2, GPR146, NECTIN4, PIK3R1, SPON1, SPAG4, RAPGEF3, IQGAP2, BHLHE40, CEBPA, PREP, CORO2A, ANKRD37, ZNF555, P4HA1, PLIN2 |
| PE with FGR | Late-onset, mild-to-moderate | Endometrial maturation | Early-secretory phase | MID1, MT1F, TIMP1, ANKRD37, PKM, ACVR1, ISG15, IFIT1, TP53INP1, HILPDA, CITED2, TM9SF3, TMEM106B, SFRP1, FHL1, RPS6KA6, POGK, SGMS2, KRT17, TIPARP, TMSB10, PTPRE, PARP12, BPHL, ARL6IP5, STARD8, CLOCK, FAM107B, PPP1R14C, LAMC2, KRT19, CHI3L1, PTGFRN, CLIC5, TLR3, SPAG4, PAPSS1 |
| PE with FGR | Late-onset, severe | Endometrial maturation | Early-secretory phase | CDH11, PJA1, DESI2, IFITM3, MMD, PMP22, PKD2, LTBP1, AK4, CHMP2A, CHSY1, SEPHS1, SIK3, STX6, PMAIP1, HPCAL1, FBN2, RASSF8, FBLN5, MIIP, VIM, LEPROTL1, MEIS1, C4orf19, PDS5B, APBB2, FXYD5, SESN3, PROCR, RPS6KA5, ATP7B, QPCT, MTFMT, SOWAHC, SYDE1, HEY1, SSR1, NFKB1, DIO2, PROM1, SPON1, PDGFC, LSM6, CORO1C, RABEP1, GTDC1, RNF20, PALLD, PARVA, PRRX1, HMGB3, LYN, INHBB, APLN, RAB8B, FHL2, MME, PAWR, THOP1, ESYT2, ARID3A, SERPINE1, NCEH1, INPP5F, CEBPG, SHANK2, TBX3, SPAG4, CH25H, UBE2Q2, GYPC, TPM2, SLC6A8, MARCKS, CDYL2, LRRC1, PKP2, SH3BP5, OPN3, ZDHHC14, ING3, SYK, MGAT5, RCBTB2, KLHL3, PKNOX1, DSTYK, FSTL3, NFIA, PARM1, SNX18, ATP1A1, MYO5A, LAMA4, PPP2R5E, SEMA3G, EPHB3, CDO1, SLC36A1, OLFML2B, ZBTB2, CORO1A, ARHGDIB, NPR3, PRKAB2, SLC25A19, DAB2IP, PHTF2, WNT5A, TSC22D2, STX1A, TAF9B, MMP2, RIOK1, TNFRSF10B, CREB1, DDIT4, HECW2, RNF130, ZNF614, PHYHIPL, CYFIP2, BHLHE40, USP46, PPP2R3A, TNFRSF10D, NFIB, GULP1, CST3, EFNB1, GOT1, CALM1, MAD2L2, GIMAP4, CRY1, ZNF350, SPRY1, TNFAIP8, CPEB4, ABAT, NDUFB9, GRINA, CDC42BPA, COG2, PTPRD, ARSA, RNF19A, DLG3, ADM, CLDN3, STX12, MFGE8, RDH10, GORAB, GPR146, SPRED1, SEC61B, BASP1, RNF114, DUSP1, FAM107B, RHEB, AKAP13, DDA1, ABHD5, DDHD1, PKM, PTPRJ, TRIM25, VEGFA, EPB41L4B, SLC8B1, GRHL1, SUSD1, ARRDC3, SUV39H1, SLCO2A1, CLIC5, EZR, SH3RF1, KLF3, PXK, LRRC42, IQGAP2, IGSF3, PLEKHF2, DDR2, GPER1, DYNLL1, RNF24, SH3PXD2A, OPRK1, ANGPTL2, ZEB2, AASDHPPT, C1orf21, UBALD2, RARRES1, HTRA1, NEDD4L, GPX3, SLCO4A1, PREP, EVI5, MFSD1, NUFIP2, MID1, WDSUB1, CIB2, DSP, CALM3, BCAR3, SLIT3, ERCC6, CCND3, KITLG, NECTIN4, LRIG3, PTBP3, CYBRD1, REV3L, NRIP1, EFNA1, KRT19, FPGS, APOL4, SLC23A2, ACAA2, CALCOCO1, TACC2, DOCK5, TSPAN31, PHF19, MOB3C, DNMT3A, PGBD2, RERG, ARL6IP5, MATN2, GNAQ, FLNB, EAF1, UROS, TMCC3, MBIP, TBL1XR1, ARRDC4, NCAPD2, FAM120AOS, CITED2, DCP2, LEP, SLC44A3, RUNX1T1, LURAP1L, SNAPC1, NAXD, SNAI2, BPGM, DONSON, MORC3, FSCN1, NEK11, LRRC41, AHNAK, PFKFB3, BGN, S100A6, WDR11, TXNRD1, SQLE, SOCS2, RASEF, MXI1, IGFBP4, PLPP3, PDE9A, ENG, RNASE1, DUSP4, HNRNPA0, SLC17A5, BTD, CX3CR1, TM4SF1, ICA1, TXNIP, GCLM, FAM210A, CA2, MYO9A, ERRFI1, PIK3CB, ANGPT1, SERPINB9, IFT20, ZC3H6, CYB561A3, TIMP4, MAPRE2, TBC1D4, PRKCH, CA4, CYP2J2, PRRG4, TM9SF3, FOSL2, CTBS, CRYAB, ESRP1, LAMB1, COL1A2, CEP68, P4HA1, ACTG2, SLC2A1, MSH6, TLR4, SWAP70, ADAMTS5, EIF4G2, SCARB1, SGK1, PLOD2, HILPDA, VPS25, TGFBR3, DISP1, SERTAD2, PLIN2, DLX5, PAPPA, ADCY7, CNOT8, FRMD4A, NRBF2, RIN2, NFIL3, CMYA5, ERP44, FBXO28, MAP3K8, CCSAP, BCAT2, GNG11, SLC12A6, GPT2, PPP1CB, RAPH1, PDGFRA, TPD52L1, VEGFC, MPC1, TACC1, ANKRD37, MAOA, SEC11C, SLC30A2, PPL, BTAF1 |
|  |  |  | Late-secretory phase | VPS45, GSTK1, COL6A1, SERPINB9, PALMD, FXYD5, MATN2, CCDC113, PTPRJ, RDH10, SUV39H1, GBE1, KLF3, SARAF, CLCN5, GNAQ, PDGFRA, RUNX1T1, LSM6, TNFAIP8L1, DNMT3A, REV3L, ZFP3, DUSP1, SWAP70, DIAPH2, ACKR2, SHANK2, FAM107B, CITED2, C3AR1, TM4SF1, MORC3, RNF20, NVL, EFNA1, RASEF, COIL, INHBB, RGS16, RNF19A, LRRC41, ZNF75D, FGD6, SH3BP5, UBALD2, TRIM24, RIN2, TMEM176B, IVD, CD163, JMY, TBXAS1, ELL, ASAP2, CTBS, CASP4, UBR1, WWTR1, NCAPD2, TPM2, IGSF3, LYN, AMPD3, TMCC3, SMARCA2, NR4A3, DYNLL1, SQLE, IRF2BPL, PTBP3, CRY1, CYBRD1, NFIL3, TRIM25, DEGS1, SLC25A4, WDSUB1, PAM, CDC42BPA, STK17A, GRHL1, CLIC3, TNFAIP8, TAF9B, MFSD1, BTAF1, ABTB2, IGSF8, GRINA, SLC23A2, QPCT, DNAJB9, CDO1, UROS, DDA1, PAPPA, GORAB, CALM1, COL17A1, RNASE1, EPS8L1, PKNOX1, ELF4, RFK, TSC22D2, PROM1, ZFAND5, NCEH1, ZFP36L1, SLC25A19, SH3BGRL, NR2F1, MAML2, HMGN3, SLC17A5, SESN3, ANTXR2, PKM, DIP2B, NBAS, ARHGEF12, MME, BHLHE40, RARRES1, ERO1A, NRBF2, SLC16A3, MAD2L2, AHNAK, HTRA1, TIMMDC1, ASTE1, TSC22D1, EAF1, HNRNPA0, ELMO1, ST6GAL1, CYBB, INPP5F, HECW2, WFS1, CORO2A, CALD1, GPER1, FNDC3A, TMEM140, RNF24, FHL2, GULP1, KIF17, LTBP1, AZIN1, PHYHIPL, CIB2, C1QC, PLIN2, CYFIP2, NFIA, PPP2R5A, SPON1, VIM, DONSON, KAT14, PKP2, IGFBP4, SEC11C, FAM43A, SLC8B1, PDGFC, HPGDS, ESYT2, VEGFA, HEY1, EPHB3, MTMR6, CORO1C, RPS6KA5, COPZ2, C4orf3, ZNF614, SEPHS1, MAOA, ADM, TTLL5, ENG, SGCB, MMP2, DCN, TACSTD2, SOAT1, C1QB, UPP1, LAMB1, SH2D4A, FRMD4A, SYNJ2, SLC9A9, PIK3R1, CA2, DLX5, DDR2, IGFBP5, PJA1, MANBA, SLC2A1, LURAP1L, PGBD2, NDUFA4, SERPINE1, CDYL2, MSH6, PCM1, THOP1, CALM3, ABHD5, TPD52L1, SLCO2B1, G6PC3, FBLN5, GATA3, SOWAHC, BAD, SPAG4, IGF1, SLC12A6, MOCOS, MOB3C, NAXD, HILPDA, MREG, CRYAB, BCAT2, RDX, BCAR3, CX3CR1, TREM1, SERPINB2, NRIP1, DUSP4, SLC30A2, PALLD, ZDHHC14, GREM2, SH3PXD2A, PIGX, RAB8B, STAB1, CHSY1, PDS5B, DSTYK, IFITM3, CLIC5, SLCO4A1, ERLIN1, EFHD1, PMP22, MXI1, ARHGDIB, PPL, GPX3, HMGB3, FAM172A, FAM210A, NDUFB9, PRRG4, SEMA4B, GALNT10, MID1, COL1A2, ARRDC3, FBN2, AK4, GIMAP4, ANKRD37, AKAP13, ERP44, DDIT4, CFL2, RAPH1, PREP, LRRC8A, MYO5A, SNAPC1, PFKFB3, CORO1A, SLC1A1, BPGM, PIK3CB, BAZ1A, PIK3CG, SGK1, F3, PLOD2, TYROBP, MAP3K8, GLUL, FOSL2, PIK3AP1, ADAMTS5, PRKCH, LCP1, CPEB4, TBC1D15 |
|  |  | Placenta | First trimester | C1orf21, UPP1, AZIN1, ATP7B, CCDC113, DIP2B, MLLT10, ACSF2, ARL6IP5, SH2D4A, PIK3AP1, TLR4, SLC30A2, MME, CDO1, HIC2, PRKCA, MAN1A2, SLC36A1, CH25H, NRBF2, CCSAP, ANKDD1A, PFKFB4, TRMT6, USP46, WDSUB1, WEE1, SNAPC1, NPEPPS, TACSTD2, CTBS, TPM2, HECW2, GNG11, UROS, KLHL3, NCEH1, NEDD4L, BTAF1, TACC1, SLC44A3, SPON1, DDR2, STAT5B, THOP1, IFITM3, GRHL1, INSL4, ARRDC3, IGFBP4, SEMA3G, VEGFA, EPS8L1, GMFB, SOWAHC, NFIA, SLC11A1, GCLM, DOCK5, CYBB, TMEM140, MYO5A, DYRK2, KAT14, F8, ADCY7, SEMA4C, AKAP13, STX6, ANGPTL2, C3AR1, RCBTB2, PRKCE, SLC2A1, GRHL3, PLOD2, INPP5F, MAN1C1, PRKCH, RBM43, DLX5, MAP3K9, PREP, ZNF331, RASSF8, FNDC3A, FBN2, SLC35F6, DUS2, SQLE, TBC1D15, ZNF75A, SLCO2B1, SLC8B1, GNAQ, DDIT4, SH3RF1, CMKLR1, PKM, ACO1, INHBB, GATA3, SLIT3, BGN, RNF146, OLFML2B, TDP1, RNF130, RHEB, MSH6, ANGPT1, TIMP4, APLN, PROM1, NPL, PPP2R5E, KRT4, UBE2H, SLC6A8, PMAIP1, KIAA0040, MATN2, ESRP1, HK1, PDP2, UTP15, CMYA5, ARRDC4, CALM3, RDH10, MCTP2, SCARB1, NFIB, ABTB2, JAK2, LRRC8A, TTI1, CA8, ERRFI1, GALK1, BRAF, TSC22D2, NECTIN4, CA4, KIF2A, PDE9A, MTMR4, PARM1, CSNK2A2, ING3, EIF4G2, EPB41L4B, MKNK1, VEGFC, MFGE8, MIIP, PCM1, SLC16A3, LIG3, DUSP4, WDR11, TM4SF1, PKD2, SNAI2, AGTR1, INSIG1, SESN3, HMGN3, PPP2R5A, RLF, MREG, GSE1, LEPROTL1, NPR3, SOCS2, GRINA, RANBP10, AP5B1, PDGFC, COL6A1, CDH11, SYDE1, NGLY1, MMP2, RAB3B, RASEF, BTD, C1QB, KIF17, IRF2BPL, MXI1, IRX2, SARAF, IGFBP5, TSPAN5, KIF5B, TMCC3, YTHDC2, LURAP1L, SLC6A4, CLIC5, ASTE1, EFHD1, KCNK5, BCORL1, AASDHPPT, ANTXR2, NR2F1, RARRES1, MOB3C, TMTC4, RIN2, PALMD, LIMCH1, RIMKLA, AVL9, CHSY1, ELMO1, C4orf19, PAPPA, VIM, ST6GAL1, SMARCA2, DIO2, HEY1, ARID3A, ZFP36L1, PIK3CB, RNF20, PRKAB2, SHANK2, ASAP2, ACTG2, KITLG, PDE5A, LAMA4, NFKB1, CD200, ADARB1, LTF, CDYL2, GALNT18, DAB2, PLA2G5, IGF1, KMT2E, TMEM176B, TLR7, CADM3, PTPRJ, CPOX, GPT2, DONSON, SPRY1, WWTR1, SHE, GIMAP4, COIL, TBC1D4, SLC9A9, TYROBP, TRIP10, WNK2, GAN, SYK, RNASE1, C1QC, RRAS, TBXAS1, TXNIP, CLDN5, MAPRE2, RPS6KA5, MAML2, PFKFB3, GPER1, CX3CR1, DUSP1, WWC3 |
|  |  | Placenta | Second trimester | SYDE1, RAB3B, SCARB1, CDK19, DUSP4, NECTIN4, SLC35F2, PAPPA2, APLN, CLIC5, TMEM140, EFHD1, ENG, PLCXD2, PLOD2, MAN1A2, C1orf21, MAN1C1, SERPINE1, GPER1, LIMCH1, TIMP4, CD200, PAM, KIF2A, PAPPA, UROS, HEY1, CCSAP, ACSF2, SOWAHC, PRKCA, EPS8L1, STAT5B, SNAI2, CDH11, C4orf19, HMGN3, TMTC4, PRKCH, MTMR4 |
|  |  | Chorioamnionitis | Second trimester | ERRFI1, LIMCH1, CX3CR1, QPCT, LRRC1, RASEF, INHA, GPT2, DLX5, PROCR, FSTL3, PIK3AP1, NEK11, CCDC113, SH3PXD2A, SLCO2A1, LEP, KRT19, SYDE1, ENG |
|  |  |  | Third trimester | MAP3K9, SLC16A3, PPP1R12C, IRF2BPL, CLDN5, ACKR2, TSC22D1, FAM219A, DAB2IP, PGBD2, GBE1, CCDC113, C4orf3, GALK1, TSC22D2, BCAT2, RPS6KA5, PLEC, STX1A, EPS8L1, SLC35F2, SMAD2, UBR1, MAN1C1, RIOK1, CCDC47, UBE2Q2, GSE1, HIC2, RLF, ELL, RANBP10, MARCKS, PURB, FOXJ2, RIMKLA, BTD, DESI2, AK4, ARRDC3, GATA3, CA4, ZNF350, CLIC3, FGD6, RHEB, S100A6, NVL, SLC6A8, DIO2, PPP2R5E, PJA1, BAD, ARID3A, DSP, TNFRSF10B, CRKL, MOB3A, PDS5B, LEPROTL1, SCARB1, ZC3H6, TACC1, PPL, P4HA1, PIK3R1, GPR146, ANXA4, ASTE1, SLC28A1, PFKFB3, KIF17, CDK19, CLDN3, MAML2, TMEM45A, TRIP10, SARAF, LRRC8A, SHANK2, P2RY2, FBXO28, SEMA4C, ESYT2, PROCR, PCM1, MORC3, SP1, TRIM24, CSNK2A2, SYNJ2, ANKRD37, TNFRSF10D, IGSF3, LTBP1, PAM, ENG, ZNF175, BTAF1, MID1, GAN, SYN3, RARRES1, SSR1, STK24, CRY1, CAPN7, ERP44, SOWAHC, QPCT, PPP2R5A, SNX18, EVI5, CEBPG, CLCN5, CMYA5, SLC11A2, GPER1, SEMA3G, MSH6, PRRG4, RNF24, PAPPA2, SLC17A5, VIM, VEGFC, WDR11, FSTL3, HPCAL1, FBLN5, NRIP1, GYPC, SLCO2A1, SIGLEC6, F3, PMP22, STAB1, GREM2, COPZ2, ERRFI1, MRPS30, INHA, PIGX, ZC2HC1A, PKNOX1, CALD1, SERTAD2, PKP2, SUV39H1, RBM22, FBN2, TREM1, IFITM3, CD4, ENY2, PHF19, EFHD1, HEY1, APBB2, ZMYND8, SLC11A1, SYK, LTF, ZNF555, SPAG4, CORO1A, GRHL3, TACC2, PARVA, MAP3K8, LAMB1, ERLIN1, AVL9, MANBA, ALKBH5, CX3CR1, HILPDA, SH3BP5, C1QC, COL17A1, TNFAIP8L1, MTMR4, KIF2A, PDE9A, F8, CALM3, SLC30A2, FAM107B, FAM20B, ERO1A, EAF1, BCAR3, PAWR, SQLE, SLCO2B1, MKNK1, FOXJ3, SLC6A4, CD163, ATP7B, TPM2, SGK1, SNAI2, SYDE1, WWTR1, COL6A1, MXI1, TTLL5, NDUFA4, TNFAIP8, ADAMTS5, SDF2L1, ING3, ZBTB2, MMP2, RNF130, ELMO1, COL1A2, SH2D4A, HTRA1, PALLD, RERG, NR2F1, ANGPTL2, LEP, PIK3CB, MLLT10, GNG11, CFL2, STAT5B, ICA1, PLIN2, CCDC127, SOCS2, SFXN3, CDO1, COG3, PDGFRA, DCP2, LYN, PLPP3, SLC2A1, MGAT5, SERPINB9, SPON1, TSPAN5, SLC25A25, CASP4, NEK11, WWC3, GPT2, OLFML2B, CTSZ, APOL4, SLCO4A1, CYBRD1, DNMT3A, AASDHPPT, WFS1, FSCN1, SLIT3, TDP1, RCBTB2, KRT19, FAM120AOS, SH3PXD2A, DUSP1, PRKCH, HMGB3, APLN, PMAIP1, LRRC42, ANTXR2, RASSF8, CHMP2B, C3AR1, SEPHS1, CDH11, DLX5, CEBPA, ACTG2, OPN3, PRRX1, NECTIN4, GOT1, PTBP3, RNF20, USP46, EFNB1, DCN, TYROBP, LRRC1, GFOD2, RFK, BHLHE40, PIK3AP1, RDH10, UBE2H, CALM1, MTFMT, GRHL1, CD200, IGF1, SEC22C, YTHDC2, LIMCH1, EZR, CREB1, PTPN1, AKAP13, FAP, SH3BGRL, GSTK1, ADCY7, ARL2BP, SMARCA2, ZFAND5, PRKCA, RASEF, ARHGDIB, NAXD, VPS45, SAR1A, CORO2A, ARMCX1, SLC35F6, NDUFB4, ACO1, CCSAP, PHYHIPL, INSIG1, KMT2E, GRINA, RNASE1, RUNX1T1, CHMP2A, NFIA, PTPRD, PROM1, IVD, NUFIP2, GLUL, CALM2, SEC61B, PDGFC, CADM3, CDC42BPA, CD28, GTDC1, SGCB, ST6GAL1, UROS, CMKLR1, NDUFB9, KITLG, DISP1, ASAH1, TLR7, MCTP2, MYO5A, MATN2, LURAP1L, PXK, CYBB, NPL, PIK3CG, MEIS1, PKD2, TBXAS1, ZEB2, HPGDS, NEDD4L, RAPH1, SPRED1, SLC9A9 |
|  |  | HELLP syndrome | Third trimester | INHA, ENG, SH3BP5, LRRC1, GALK1, SH3PXD2A, KMT2E, FLNB, PRKAB2, PFKFB4, HTRA1, SFXN3, SYDE1, EPS8L1, HEY1, PKD2, TRIM24, MOB3A, NECTIN4, PDE5A, ING3, PAM, KLF3, GBE1, SUV39H1, ACKR2, PLCXD2, PIGX, FOSL2, PKM, TREM1, RASEF, STK24, ALKBH5, PLEC, CRKL, ELF4, TAF9B, GPX3, MIIP, ZNF331, ARID3A, EAF1, PAPPA2, PIK3R1, HK1, TRIP10, DONSON, BHLHE40, GREM2, NFIL3, PRKCH, MOB3C, ASTE1, SLC2A1, CEBPA, SLC16A3, PHTF2, BAD, PGBD2, CYB561A3, CYBB, LRRC8A, RPS6KA5, DDR2, VEGFA, DYRK2, FPGS, COIL, SH3RF1, BTD, GCLM, CCDC127, CSNK2A2, SHANK2, CALM1, DIO2, MAP3K8, HPGDS, TM9SF3, PAWR, GRHL3, GORAB, PIK3CB, PPP1R12C, PJA1, VPS45, BCAR3, FRMD4A, SLC25A4, ZC2HC1A, BASP1, SEMA4B, QPCT, PLOD2, APLN, PREP, ERO1A, TXNRD1, MYOF, SYN3, WFS1, IL36RN, CLIC3, ARRDC3, SEMA4C, ERCC6, FBN2, NOD1, WNK2, HMGB3, IQGAP2, P2RY2, SPAG4, ZNF75A, SLC6A8, SPON1, MME, SCARB1, SEPHS1, IGSF8, CREB1, NEK11, GMFB, KIT, EPHB3, FSTL3, CLDN16, SLC23A2, DLX5, IGF1, DUSP1, ZNF555, BCORL1, ERRFI1, RNF20, SWAP70, GPT2, C4orf3, VEGFC, AZIN1, BCAT2, MAML2, PROCR, P4HA1, MAP3K9, CORO2A, IGSF3, ANKRD37, SERPINB2, SPRED1, CCSAP, NPEPPS, EFHD1, FAM20B, UBE2Q2, DAB2IP, CYP2J2, AFMID, ZEB2, ASAH1, TMEM45A, TSC22D1, CD28, HPCAL1, ZMYND8, APBB2, IRF2BPL, FAM43A, GPR146, MRPS30, NUFIP2, MTMR6, CDO1, PLIN2, EFNB1, TLR7, LYN, PPL, KIF5B, LIMCH1, GATA3, PIK3AP1, KRT19, EZR, PLPP3, GRHL1, WDR11, LEP |

FGR, fetal growth restriction; HELLP, hemolysis, elevated liver enzyme, and low platelet; PE, preeclampsia.

Table S2. Discovery of gene set function by GSAI for late-onset, severe PE.

| Representation of | | Ontology | GSAI |  |
| --- | --- | --- | --- | --- |
|  |  |  | Query | Response* |
| Endometrial maturation | Mid-secretory phase | CliXO:111 | MRPL34, DYNLL1 | Process: Mitochondrial Ribosome Biogenesis and Cellular Respiration (0.85)   1. MRPL34 [D/U], or Mitochondrial Ribosomal Protein L34, is a component of the large subunit of the mitochondrial ribosome. Ribosomes are responsible for protein synthesis, and in the case of mitochondrial ribosomes, they synthesize proteins that are essential for the mitochondrial electron transport chain, which is the primary mechanism of ATP production in the cell. Therefore, MRPL34 [D/U] plays a crucial role in energy production and cellular respiration. 2. DYNLL1 [D/U], or Dynein Light Chain 1, is a motor protein involved in various cellular processes, including intracellular transport and cell division. However, it also interacts with the mitochondrial outer membrane and is implicated in the regulation of mitochondrial morphology and function. This suggests a role in cellular respiration, as the mitochondria are the site of oxidative phosphorylation, the final stage of cellular respiration.   In summary, the proteins in this system are involved in the biogenesis of the mitochondrial ribosome and the regulation of mitochondrial function, both of which are essential for cellular respiration. The interplay between these proteins likely ensures the efficient production of ATP, the cell's main energy currency. The synergistic effect of these proteins contributes to the maintenance of cellular energy homeostasis. |
| Chorio-amnionitis | Third trimester | root | ANKRD13A, BHLHE40 | Process: Regulation of Protein Ubiquitination and Circadian Rhythm (0.85)   1. ANKRD13A [D/D], or Ankyrin Repeat Domain 13A, is a protein that is known to interact with ubiquitin and plays a significant role in the process of protein ubiquitination. Ubiquitination is a post-translational modification that regulates protein degradation, cellular localization, and activity. ANKRD13A [D/D] is involved in the endocytic pathway, which is responsible for the internalization of proteins from the cell surface. It is also known to interact with the E3 ubiquitin-protein ligase UBR4, suggesting a role in the ubiquitin-protease system. 2. BHLHE40 [D/U], also known as Basic Helix-Loop-Helix Family Member E40, is a transcription factor that is involved in the regulation of circadian rhythm. It is a part of the negative feedback loop in the circadian clock, where it represses the activity of CLOCK-ARNTL/BMAL1 heterodimers. BHLHE40 [D/U] also plays a role in the regulation of gene expression in response to hypoxia and is involved in cellular response to stress.   In summary, the proteins in this system are involved in the regulation of protein ubiquitination and circadian rhythm. ANKRD13A [D/D] contributes to the process of protein ubiquitination, which is crucial for protein degradation and cellular localization. BHLHE40 [D/U], on the other hand, is involved in the regulation of circadian rhythm, contributing to the maintenance of the body's internal clock. The interplay between these proteins could potentially influence the balance between protein degradation and synthesis, as well as the synchronization of cellular processes with the circadian rhythm. |
|  |  | CliXO:86 | ANKRD13A, SLA | Process: Protein Ubiquitination and Immune Response Regulation (0.85)   1. ANKRD13A [D/D], also known as Ankyrin Repeat Domain 13A, is a protein that has been found to be involved in the process of ubiquitination, a post-translational modification that regulates protein degradation, cellular localization, and activity. ANKRD13A [D/D] has been shown to interact with ubiquitin and facilitate the transport of ubiquitinated proteins to the proteasome for degradation. This process is crucial for maintaining protein homeostasis within the cell. 2. SLA [U/U], Src-like-adaptor, is a protein that plays a significant role in immune response regulation. It acts as a negative regulator of T-cell receptor (TCR) signaling, which is a critical pathway in the immune response. By inhibiting TCR signaling, SLA [U/U] helps to prevent overactive immune responses and maintain immune system balance.   In summary, the proteins in this system are primarily involved in the regulation of protein ubiquitination and immune response. ANKRD13A [D/D] contributes to the maintenance of protein homeostasis through its role in protein ubiquitination and degradation, while SLA [U/U] plays a crucial role in immune response regulation through its inhibitory effect on TCR signaling. These processes are essential for maintaining cellular function and immune system balance. |
|  |  | CliXO:78 | IGF1 | Process: Insulin-like Growth Factor Signaling (1.00)  The protein IGF1 [U/D], or Insulin-like Growth Factor 1, is a hormone that plays a crucial role in cell proliferation, growth, and apoptosis. It is part of the Insulin-like Growth Factor (IGF) signaling system, which is a complex network of receptors, binding proteins, and interacting proteins.  IGF1 [U/D] is primarily produced in the liver as an endocrine hormone and plays a significant role in childhood growth and continues to have anabolic effects in adults. It is structurally similar to insulin and can even bind to the insulin receptor, albeit with lower affinity than insulin itself.  The IGF1 [U/D] receptor (IGF1 [U/D] R) is a transmembrane receptor that is activated by IGF1 [U/D]. Upon activation, the receptor undergoes autophosphorylation and triggers a cascade of intracellular signaling pathways, including the PI3K-AKT and the MAPK pathways. These pathways regulate various cellular processes such as cell growth, differentiation, and survival.  IGF1 [U/D] also interacts with a family of six IGF-binding proteins (IGFBP1-6) that regulate its biological activities. These binding proteins can prolong the half-life of IGF1 [U/D], inhibit its interaction with IGF1 [U/D] R, and modulate its endocrine, paracrine, and autocrine actions.  In summary, the IGF1 [U/D] protein is a key player in the IGF signaling system, which is a complex network of interactions that regulate cell growth, differentiation, and survival. The system exhibits both synergistic and antagonistic effects, depending on the context and the specific interactions involved. |
| HELLP syndrome | Third trimester | CliXO:70 | PDE5A | Process: Cyclic Guanosine Monophosphate (cGMP) Degradation (0.95)  PDE5A [D/D], or Phosphodiesterase 5A, is a key enzyme involved in the degradation of cyclic guanosine monophosphate (cGMP), a secondary messenger molecule that plays a crucial role in various physiological processes.  The primary function of PDE5A [D/D] is to hydrolyze cGMP to 5'-GMP, thereby terminating its signaling function. This protein is predominantly found in vascular smooth muscle cells, where it plays a critical role in regulating vascular tone and blood flow. By degrading cGMP, PDE5A [D/D] modulates the effects of nitric oxide (NO), a potent vasodilator that increases the concentration of cGMP in cells.  In the context of the cardiovascular system, PDE5A [D/D]'s activity is crucial for the regulation of blood pressure and cardiac function. Inhibition of PDE5A [D/D], as seen with drugs like sildenafil, can enhance the vasodilatory effects of NO, leading to decreased blood pressure and improved blood flow.  In the nervous system, PDE5A [D/D] also plays a role in the regulation of neuronal signaling. cGMP is involved in the modulation of synaptic plasticity and memory formation, and the activity of PDE5A [D/D] can influence these processes.  In summary, the primary biological process performed by PDE5A [D/D] is the degradation of cGMP, a process that has significant implications for vascular and neuronal function. The protein's role in this process is well-established, hence the high confidence score. |
|  |  | CliXO:71 | FAM43A | Without additional proteins to analyze, it's impossible to provide a comprehensive analysis of the biological processes performed by this system of interacting proteins. FAM43A [U/U] alone is not sufficient to determine a prominent biological process.  Process: System of unrelated proteins (0.00)  FAM43A [U/U], also known as Family with sequence similarity 43 member A, is a protein that in humans is encoded by the FAM43A [U/U] gene. The function of this protein is not well understood, but it is believed to play a role in cellular processes due to its ubiquitous expression in various tissues. However, without additional proteins to interact with, it's impossible to determine a specific biological process in which FAM43A [U/U] is involved.  In conclusion, without additional proteins for analysis, the system can only be identified as a "System of unrelated proteins" with a confidence score of 0.00. Further research and data are needed to elucidate the specific biological processes and interactions involving FAM43A [U/U]. |

*, confidence score is shown in parentheses after the process name and each mention of gene name is modified by including up (U)- or down (D)-regulation status of respectively the associated factor and third-trimester placenta under late-onset, severe PE; CliXO, clique-extracted ontology; GSAI, gene set artificial intelligence; HELLP, hemolysis, elevated liver enzyme, and low platelet; PE, preeclampsia.

Table S3. Discovery of gene set function by GSAI for early-onset, severe PE.

| Representation of | | Ontology | GSAI |  |
| --- | --- | --- | --- | --- |
|  |  |  | Query | Response* |
| Endometrial maturation | Late-secretory phase | CliXO:449 | SNAP23, LDLR, PPL, PTBP3, LRRC32 | Process: Lipid Metabolism and Cellular Adhesion (0.85)   1. SNAP23 [U/D] is a member of the SNARE family of proteins, which are involved in vesicle trafficking processes. Specifically, SNAP23 [U/D] is involved in the docking and fusion of vesicles to target membranes, a process that is crucial for the secretion of lipoproteins and other molecules. 2. LDLR [U/U] is a receptor protein that binds low-density lipoprotein (LDL), the primary cholesterol-carrying lipoprotein of plasma, and transports it into cells via endocytosis. This process is a key mechanism for the regulation of cholesterol levels in the body. 3. PPL [U/U], or Periplakin, is a component of the cornified cell envelope of keratinocytes, a structure that provides resistance to mechanical stress. It also plays a role in cellular adhesion and the organization of intercellular junctions. 4. PTBP3 [U/U], also known as ROD1, is a regulator of differentiation 1. It is involved in the regulation of alternative splicing, a process that allows for the generation of multiple protein isoforms from a single gene. This protein has been implicated in the regulation of lipid metabolism, particularly in the liver. 5. LRRC32 [U/U], also known as GARP, is a transmembrane protein that is expressed on the surface of regulatory T cells (Tregs). It plays a role in the suppression of the immune response, and it is also involved in cellular adhesion.   In summary, the proteins in this system are involved in lipid metabolism, cellular adhesion, and the regulation of the immune response. The interplay between these proteins allows for the regulation of cholesterol levels, the maintenance of cellular structure and integrity, and the modulation of the immune response. The proteins in this system work together to ensure the proper functioning of these processes, with potential synergistic effects in lipid metabolism and cellular adhesion. |
| Placentation | First trimester | CliXO:410 | GPT2, UBE2H, NIPAL3 | Process: Glutamate Metabolism and Protein Ubiquitination (0.85)   1. GPT2 [U/U], also known as Glutamic Pyruvate Transaminase 2, is a key enzyme involved in the metabolism of amino acids, specifically the conversion of alanine and alpha-ketoglutarate to pyruvate and glutamate. This process is crucial for the regulation of energy production and the maintenance of nitrogen balance in the body. 2. UBE2H [U/D], or Ubiquitin-Conjugating Enzyme E2 H, is a member of the ubiquitin-conjugating enzyme family. It plays a significant role in the ubiquitin-proteasome system, which is responsible for the degradation of intracellular proteins. This process is essential for maintaining cellular homeostasis by removing misfolded or damaged proteins, controlling the levels of certain proteins, and regulating various cellular processes. 3. NIPAL3 [U/D], also known as Magnesium Transporter NIPA3, is involved in the transport of magnesium, a critical cofactor for many enzymes, across membranes. However, recent studies have also suggested a role in lipid metabolism, particularly in the formation and maintenance of the epidermal barrier.   In summary, the proteins in this system are primarily involved in the metabolism of amino acids, protein ubiquitination, and potentially lipid metabolism. These processes are interconnected, as the degradation of proteins (mediated by UBE2H [U/D]) can provide amino acids for further metabolism (mediated by GPT2 [U/U]), and both processes could potentially interact with lipid metabolism (mediated by NIPAL3 [U/D]). The interplay between these proteins and their associated processes contributes to the maintenance of cellular homeostasis. |
|  |  | CliXO:417 | GPT2, NIPAL3, FGF13, NIN, KIAA0232, MT1F, ZBTB20, DKK3, PDE5A | Process: Mitochondrial Metabolism and Neurodevelopment (0.85)  1. GPT2 [U/U] and NIPAL3 [U/D] are both involved in mitochondrial metabolism. GPT2 [U/U], or Glutamic Pyruvate Transaminase 2, is a key enzyme in the alanine pathway, which is one of the most significant gluconeogenesis pathways. It catalyzes the reversible conversion of alanine and alpha-ketoglutarate to pyruvate and glutamate, playing a crucial role in amino acid metabolism and the tricarboxylic acid cycle. NIPAL3 [U/D], or Magnesium Transporter NIPA3, is a magnesium transporter protein that is localized in the mitochondria and is essential for maintaining mitochondrial magnesium homeostasis.  2. FGF13 [D/D], NIN [D/D], and ZBTB20 [D/D] are all involved in neurodevelopment. FGF13 [D/D], or Fibroblast Growth Factor 13, is a non-secreted modulator of neuronal excitability that regulates the voltage-gated sodium channels in the brain, playing a crucial role in neuronal development and function. NIN [D/D], or Ninein, is a centrosomal protein that is essential for microtubule anchoring and spindle formation during cell division, playing a significant role in neuronal migration and differentiation. ZBTB20 [D/D], or Zinc Finger and BTB Domain Containing 20, is a transcriptional repressor that is involved in neurogenesis and the development of the hippocampus.   2. DKK3 [D/D], PDE5A [D/D], and MT1F [U/U] are involved in various cellular processes. DKK3 [D/D], or Dickkopf WNT Signaling Pathway Inhibitor 3, is involved in embryonic development through its inhibition of the WNT signaling pathway. PDE5A [D/D], or Phosphodiesterase 5A, is involved in the regulation of intracellular concentrations of cyclic nucleotides and is implicated in the regulation of vascular smooth muscle contraction. MT1F [U/U], or Metallothionein 1F, is a protein that binds heavy metals, helping to detoxify these elements. 3. KIAA0232 [D/D] is a protein with unknown function. It is predicted to be a membrane protein and may be involved in cellular transport processes.   In summary, the proteins in this system are primarily involved in mitochondrial metabolism and neurodevelopment, with some proteins also contributing to cellular detoxification, embryonic development, and vascular regulation. The interplay between these proteins likely contributes to the overall function of the system, with potential synergistic effects in energy metabolism and neurodevelopment, and antagonistic effects in cellular detoxification and development. |
|  |  | CliXO:399 | MEF2A, CX3CR1 | Process: Regulation of Immune Response and Neuronal Survival (0.85)  1. MEF2A [D/U] (Myocyte Enhancer Factor 2A) is a transcription factor that plays a crucial role in the regulation of gene expression in neurons and muscle cells. In neurons, MEF2A [D/U] is involved in neuronal survival, differentiation, and synaptic plasticity. It is also known to regulate the expression of several genes involved in the immune response, including those involved in T-cell activation and differentiation.  2. CX3CR1 [D/D] (C-X3-C Motif Chemokine Receptor 1) is a receptor for the chemokine fractalkine (CX3CL1). It is expressed on the surface of various immune cells, including monocytes, macrophages, and T cells, and plays a key role in the migration and adhesion of these cells. In the central nervous system, CX3CR1 [D/D] is expressed on microglia, the resident immune cells of the brain, and is involved in neuroinflammation and neurodegeneration.  The interaction between MEF2A [D/U] and CX3CR1 [D/D] suggests a complex interplay between the immune system and the nervous system. MEF2A [D/U], through its role in gene regulation, could potentially influence the expression of CX3CR1 [D/D] and thus modulate immune responses. Conversely, signals received through CX3CR1 [D/D] could impact the activity of MEF2A [D/U], influencing neuronal survival and function. This system of interacting proteins, therefore, appears to be involved in the regulation of immune responses and neuronal survival, with potential implications for diseases involving neuroinflammation and neurodegeneration. |
|  | Second trimester | root | PAPPA2, SLC24A3 | Process: Regulation of Extracellular Matrix Remodeling and Ion Transport (0.85)   1. PAPPA2 [D/U], or Pappalysin-2, is a metalloproteinase that specifically cleaves IGFBP-5, one of the six IGF binding proteins. By cleaving IGFBP-5, PAPPA2 [D/U] releases IGFs, which are involved in various processes such as cell growth, development, and metabolism. PAPPA2 [D/U] is also involved in the regulation of extracellular matrix (ECM) remodeling, which is crucial for various biological processes including embryogenesis, tissue repair, and disease progression. 2. SLC24A3 [U/U], or Solute Carrier Family 24 Member 3, is a potassium-dependent sodium/calcium exchanger. It is involved in maintaining the homeostasis of intracellular concentrations of sodium, potassium, and calcium ions, which is crucial for various cellular processes including signal transduction, muscle contraction, and maintaining the cell's osmotic balance.   In summary, the proteins in this system are involved in the regulation of extracellular matrix remodeling and ion transport. PAPPA2 [D/U]'s role in ECM remodeling can influence cell behavior and tissue structure, while SLC24A3 [U/U]'s role in ion transport can affect various cellular processes. These proteins may interact in a way that their functions complement each other, contributing to the overall homeostasis of the cell. |
| HELLP syndrome | Third trimester | root | ALKBH5, SWAP70, DUSP1, KLF4, GREM2, GPR146, PIP5K1B, EZR | Process: Regulation of Cellular Differentiation and Signal Transduction (0.85)  1. ALKBH5 [U/U] is a demethylase that plays a crucial role in RNA metabolism and adipogenesis. It is involved in the regulation of mRNA processing and export, contributing to the overall process of gene expression.  2. SWAP70 [U/U] and PIP5K1B [D/D] are involved in the regulation of actin cytoskeleton organization. SWAP70 [U/U] is a signaling protein that mediates the activation of RAC1, a key regulator of actin cytoskeleton dynamics. PIP5K1B [D/D], on the other hand, is a kinase that generates phosphatidylinositol 4,5-bisphosphate, a critical component of the inner cell membrane that plays a role in signal transduction and cytoskeleton organization.  3. DUSP1 [U/U] is a dual specificity phosphatase that dephosphorylates MAP kinase, thereby playing a role in the MAP kinase signal transduction pathway. It is involved in the regulation of cellular processes such as proliferation, differentiation, and stress response.  4. KLF4 [U/U] is a transcription factor that plays a key role in maintaining stem cell pluripotency and promoting cellular reprogramming and differentiation. It is involved in various biological processes, including cell cycle regulation, apoptosis, and epithelial to mesenchymal transition.  5. GREM2 [U/U] is a BMP (bone morphogenetic protein) antagonist that plays a role in regulating organogenesis and differentiation. It is involved in various developmental processes, including limb bud formation, skeletal development, and neural tube closure.  6. GPR146 [U/U] is an orphan G-protein coupled receptor that is thought to be involved in signal transduction, although its exact function is not well understood.  7. EZR [U/U] is a member of the ezrin-radixin-moesin (ERM) family of proteins that link the actin cytoskeleton to the plasma membrane. It plays a role in various cellular processes, including cell adhesion, migration, and signal transduction.  In summary, the proteins in this system are involved in various aspects of cellular differentiation and signal transduction. They contribute to the regulation of gene expression, actin cytoskeleton organization, MAP kinase signaling, stem cell pluripotency, organogenesis, and potentially other signaling pathways. Their interplay likely results in a complex network of regulatory mechanisms that control cellular differentiation and response to external signals. |

*, confidence score is shown in parentheses after the process name and each mention of gene name is modified by including up (U)- or down (D)-regulation status of respectively the associated factor and third-trimester placenta under early-onset, severe PE; CliXO, clique-extracted ontology; GSAI, gene set artificial intelligence; HELLP, hemolysis, elevated liver enzyme, and low platelet; PE, preeclampsia.

Table S4. Additional analysis after identifying FAM43A and PDE5A for late-onset, severe, PE.

| Step | Tool (configuration) | Query and response |
| --- | --- | --- |
| 1 | STRING (*Homo sapiens*; add more nodes) | Query:  FAM43A, PDE5A  Response:  <http://web.archive.org/web/20240509100709/https://string-db.org/cgi/network?taskId=bxpOJnCdjCR1&sessionId=bAyDyDpxQbxF> |
| 2 | STRING (*Homo sapiens*) | Query:  FAM43A, TMEM44, LSG1, GMPS, PDE5A  Response:  <https://web.archive.org/web/20240509101256/https://string-db.org/cgi/network?taskId=b5K2497K7IUW&sessionId=bRGt4MEYRZHy> |
| 3 | GSAI (GPT-4) | Query:  FAM43A, TMEM44, LSG1, GMPS, PDE5A  Response:  Process: Ribosome Biogenesis and Purine Metabolism (0.85)   1. FAM43A and TMEM44 are proteins with less characterized functions. However, FAM43A has been suggested to play a role in cellular processes such as cell proliferation and differentiation, while TMEM44 is a transmembrane protein, which are often involved in signal transduction and cellular communication. 2. LSG1 is a key protein involved in the late steps of ribosome biogenesis, a process that is fundamental to protein synthesis in the cell. It is involved in the maturation of the 60S ribosomal subunit, a crucial component of the ribosome. 3. GMPS is an enzyme involved in the de novo synthesis of guanine nucleotides which are critical for DNA and RNA synthesis, and also play roles in signal transduction and protein synthesis. This protein is part of the purine metabolism pathway, which is essential for cell growth and proliferation. 4. PDE5A is a member of the phosphodiesterase family of proteins, which are involved in breaking down cyclic nucleotides like cAMP and cGMP. This particular protein, PDE5A, specifically breaks down cGMP, a second messenger molecule that plays important roles in signal transduction pathways.   In summary, the proteins in this system are involved in critical cellular processes such as ribosome biogenesis (LSG1), purine metabolism (GMPS), and signal transduction (PDE5A). The roles of FAM43A and TMEM44 are less clear, but they may contribute to cell proliferation and differentiation, and cellular communication, respectively. The interplay between these proteins could potentially influence cell growth and protein synthesis, although further research would be needed to fully elucidate these interactions. |

GSAI, gene set artificial intelligence; PE, preeclampsia.
